# Sociodemographic characteristics of SARS-CoV-2 serosurveillance studies with diverse recruitment strategies, Canada, 2020 to 2023

**DOI:** 10.1101/2024.12.30.24319767

**Authors:** Matthew J. Knight, Yuan Yu, Jiacheng Chen, Sheila F. O’Brien, David L. Buckeridge, Carmen Charlton, W. Alton Russell

## Abstract

**Background:** Serological testing was a key component of severe acute respiratory syndrome coronavirus-2 (SARS-CoV-2) surveillance. Social distancing interventions, resource limitations, and the need for timely data led to serosurveillance studies using a range of recruitment strategies, which likely influenced study representativeness. Characterizing representativeness in surveillance is crucial to identify gaps in sampling coverage and to assess health inequities.

**Methods:** We retrospectively analyzed three pre-existing longitudinal cohorts, two convenience samples using residual blood, and one de novo probabilistic survey conducted in Canada between April 2020 – November 2023. We calculated study specimen counts by age, sex, urbanicity, race/ethnicity, and neighborhood deprivation quintiles. We derived a ‘representation ratio’ as a simple metric to assess generalizability to a target population and various sociodemographic strata.

**Results:** The six studies included 1,321,675 specimens. When stratifying by age group and sex, 65% of racialized minority subgroups were moderately underrepresented (representation ratio < 0.75). Representation was generally higher for older Canadians, urban neighborhoods, and neighborhoods with low material deprivation. Rural representation was highest in a study that used outpatient laboratory blood specimens. Racialized minority representation was highest in a de novo probabilistic survey cohort.

**Conclusion:** While no study had adequate representation of all subgroups, less traditional recruitment strategies were more representative of some population dimensions. Understanding demographic representativeness and barriers to recruitment are important considerations when designing population health surveillance studies.

## INTRODUCTION

Serological surveillance is a critical input to infectious disease control, including pandemic preparedness and response. In 2020, Canada launched the largest serological surveillance program in its history to monitor population immunity to severe acute respiratory syndrome coronavirus-2 (SARS-CoV-2), informing COVID-19 epidemiology and antibody dynamics. Between April 2020 and February 2021, many studies began testing blood specimens for SARS-CoV-2 antibodies [1–6]. Challenged by social distancing measures, studies used diverse strategies to recruit participants or obtain blood samples. Recruitment strategy influences study population’s characteristics and the extent to which participants represent the general population [7].

Serosurveillance studies can be broadly categorized as convenience samples, de novo probabilistic surveys, or pre-existing longitudinal cohorts. For SARS-CoV-2 serosurveillance, many countries used convenience samples of residual blood specimens due to low operational costs and ease of continued sample collection over time [1,2,8,9]. Convenience samples may introduce selection bias if certain subpopulations are excluded or poorly represented [1,8]. Probabilistic serosurveys, also deployed in many regions to monitor SARS-CoV-2, can mitigate selection biases by using stratified, weight-based approaches to recruitment. De novo designs allow tailoring recruitment to study objectives [4,5,10–12]. However, probabilistic designs are time- and resource-intensive and are sometimes limited by low response rates [6,13]. Sampling within pre-existing longitudinal cohorts can improve efficiency by leveraging an established sampling frame and study infrastructure. But this precludes tailoring the sampling frame to the current research question, and generalizability may be limited by inclusion criteria or attrition.

In this study, we introduce a simple metric for diagnosing the representativeness of subgroups within a study population. Using this metric, we evaluated the sociodemographic representativeness of six SARS-CoV-2 serosurveillance studies with diverse recruitment strategies by age, sex, race/ethnicity, urbanicity, and neighborhood measures of socioeconomic deprivation. Our findings can inform serosurveillance study design for diverse pathogens.

## METHODS

### Data

We assessed representativeness by analyzing demographic data from six Canadian study populations (Table 1). Here, we define a study to be representative if the sociodemographic composition of the study population matched the census-based target population; we make no assumptions of the sampling mechanism or inferential validity. This similarity suggests the interpretation of an effect measure may be generalizable to the target population, but does not assume the effect estimate, within an uncertainty interval, will be identical between the study and target populations [14]. The six studies included one de novo cross-sectional probabilistic sample (the Canadian COVID-19 Antibody and Health Survey 1 [CCAHS-1]), one open longitudinal cohort recruited from a marketing research panel (Action to Beat Coronavirus study [Ab-C]), two pre-existing closed longitudinal cohorts (the Canadian Longitudinal Study on Aging COVID-19 Antibody Study [CLSA], the Canadian Partnership for Tomorrow’s Health COVID-19 Antibody Study [CanPath]), and two serial cross-sectional convenience samples that used residual blood from blood donations (Canadian Blood Services [CBS]) and specimens collected for outpatient laboratory testing (Alberta Precision Laboratories [APL]). The included studies tested specimens collected from April 2020 to November 2023 with sample sizes ranging from 11,050 (CCAHS-1) to 1,039,298 (CBS). Inclusion criteria and enrollment procedures have been described previously [1,2,6,12,15,16]. This study was reported using the STrengthening the Reporting of OBservational studies in Epidemiology checklist for cross-sectional studies [17].

**Table 1:** Summary of SARS-CoV-2 serological study designs included in the study.

| Study | Design | Age | Region | Specimen type | Study time and size | De novo recruitment |
| --- | --- | --- | --- | --- | --- | --- |
| Action to Beat Coronavirus (Ab-C) | Pre-existing longitudinal open research cohort | $\geq 18$ | AB, BC, MB, NB, NL, NS, ON, PE, QC, SK, YT<sup>a</sup> | Dried blood spot | 25,110 specimens from 10,621 participants May 2020 - April 2022 | No |
| Alberta Precision Laboratories (APL) | Serial cross-sectional convenience sample | $\geq 0$ | AB | Heparinized plasma<br>Plasma<br>Serum | 210,905 specimens from 187,887 participants April 2020 - October 2022 | No |
| Canadian Blood Services (CBS) | Serial cross-sectional random sample | $\geq 18$ | AB, BC, MB, NB, NL, NS, ON, PE, SK | Serum | 1,035,580 specimens from 446,187 participants May 2020 - November 2023 | No |
| Canadian Covid-19 Antibody and Health Survey 1 (CCAHS-1) | Prospective cross-sectional cohort with direct (ages 1-24) or multi-stage (ages $\geq 25$) sampling | $\geq 1$ | AB, BC, MB, NB, NL, NT, NS, NU, ON, PE, QC, SK, YT | Dried blood spot | 11,050 specimens from 11,050 participants November 2020 - April 2021 | Yes |
| Canadian Longitudinal Study on Aging (CLSA)<sup>b</sup> | Pre-existing longitudinal closed research cohort | $\geq 51$ | AB, BC, MB, NB, NL, NS, ON, PE, QC, SK | Dried blood spot<br>Plasma | 17,310 specimens from 17,310 participants October 2020 - August 2021 | No |
| Canadian Partnership for Tomorrow's Health (CanPath)<sup>c</sup> | Pre-existing longitudinal closed research cohort | $\geq 25$ | AB, BC, MB, NB, NL, NS, ON, PE, QC | Dried blood spot | 21,720 specimens from 21,717 participants February 2021 - November 2021 | No |

Notes: <sup>a</sup>Four specimens were collected from Yukon territory and were excluded from all analyses. <sup>b</sup>Composed of comprehensive sub-cohort and tracking sub-cohort that recruited participants from seven and 10 provinces, respectively. <sup>c</sup>Composed of six distinct regional cohorts. AB: Alberta; BC: British Columbia; MB: Manitoba; NB: New Brunswick; NL: Newfoundland and Labrador; NT: Northwest Territories; NS: Nova Scotia; NU: Nunavut; ON: Ontario; PE: Prince Edward Island; QC: Quebec; SK: Saskatchewan; YT: Yukon.

From each dataset, we extracted participants’ age, sex, postal code, date of specimen collection, and self-reported race/ethnicity. We used the postal code to classify participants’ residence as urban or rural and to assign participants’ neighborhood to a quintile of the Pampalon material and social deprivation indices [18]. Material deprivation is a composite measure of education, employment, and income reflecting access to essential material resources. Social deprivation is a composite measure of living alone, single-parent families, and people who are either separated, divorced, and/or widowed, reflecting the fragility of social networks. Both measures are derived from the 2016 Canadian census [19]. We used the date of specimen collection as the sample date when available (CBS, APL); otherwise, we used the date of questionnaire completion (CanPath, CCAHS-1, CLSA) or specimen receipt (Ab-C).

Race/ethnicity information was unavailable for the APL study. Deprivation indices were available for the CBS, APL, CCAHS-1, and CLSA studies. Specimen counts for the CCAHS-1 study were rounded to base 2000 in accordance with data usage guidelines. Age was calculated as the age at specimen collection (Ab-C, APL, CBS) or questionnaire completion (CanPath, CLSA, CCAHS-1) and categorized as 0-17 years, 18-26 years, 27-36 years, 37-46 years, 47-56 years, or 57 years and older. For Ab-C specimens collected between December 2020 – April 2021 and July 2021 – September 2021, we used the 2019 baseline age since the age at current collection could not be calculated. We categorized sex as male or female and excluded participants who provided alternative responses (n = 138 [Ab-C]) from analyses involving participant sex. Because race/ethnicity data collection varied between studies and differed from census categorization, we re-classified participants as ‘white’ and ‘racialized minority’ and did not analyze specific racialized minority groups (Supplementary Tables S1-S2). For studies allowing multiple encounters with participants, we imputed missing variables when available for another encounter (CBS, Ab-C, CLSA). We classified participants who identified as both white and a racialized minority as a racialized minority, and we considered Indigenous identities as a racialized minority but conducted sensitivity analyses with different classifications. Because only CCAHS-1 collected specimens from the capital cities of the Canadian territories, we restricted our primary analysis to specimens collected from Canadian provinces but assessed territorial representativeness for CCAHS-1 separately. We excluded participants who did not meet the inclusion criteria of their respective study and who were missing age, province/territory of residence, or serology test result data. For the CBS study, we did not assess the representativeness of the 0-17-year-old age group because there were no donors younger than 17. We calculated specimen counts using complete cases within each set of demographic strata (e.g., participants missing race/ethnicity were excluded when stratifying by age, sex, and race/ethnicity but not when stratifying by age, sex, and urbanicity).

### Representation ratio analysis

To assess the representativeness of subgroups defined by one or more sociodemographic variable, we derived a *representation ratio* by dividing the proportion of study specimens in a sociodemographic subgroup by the proportion of participants in the subgroup from a target population. A representation ratio less than one indicates the group is underrepresented relative to the target population, and a ratio greater than one indicates overrepresentation. We defined the target subgroup distribution using weighted 2016 Canadian census counts [19], restricting by age and province/territory to match studies’ inclusion criteria (Table 1) and rounded to the nearest multiple of zero or five. We calculated representation ratios on unweighted study populations to assess the unadjusted sociodemographic composition of each study except for CCAHS-1, due to guidelines restricting unweighted analyses [6]. For the target distributions, race/ethnicity counts were derived using the census population group variable and Indigenous-identifying respondents were classified as racialized minorities [19]. Indigenous-identifying census respondents were classified as racialized minorities, but were excluded when generating ratios for CLSA and CanPath since Indigenous status was unavailable for most participants. We assessed the impact of excluding Indigenous-identifying census respondents in a second sensitivity analysis. We performed bootstrapping (n = 5000) to generate an uncertainty distribution for each representation ratio.

### Sample count by strata analysis

In some cases, statistical adjustment or subsampling may allow derivation of representative population statistics from large, unbalanced study populations if there are sufficient samples from less represented strata [20]. To inform whether this would be feasible in our study populations, we assessed the number of strata with counts greater than 25 when grouped by age, sex, province of residence, urbanicity, race/ethnicity, and date of specimen collection binned into two-month intervals. All analyses were conducted in R version 4.3.1 [21].

## RESULTS

### Study population

During data pre-processing, we excluded 3,718 observations for CBS (0.4%), 3,871 for APL (1.8%), 2,052 for Ab-C (7.6%), 2,024 for CLSA (10.5%), and 4,258 for CanPath (16.4%) due to missing data or failure to meet inclusion criteria. We analyzed the remaining 1,035,580 (CBS), 210,905 (APL), 25,110 (Ab-C), 21,720 (CanPath), 17,310 (CLSA), and 11,050 (CCAHS-1) observations (Supplementary Table S3). For Ab-C, CLSA, and CanPath, the minimum age of participants included in our analysis (Table 1) was older than the minimum age specified in their inclusion criteria [12,15,16]. Across studies, the largest number of observations were in the 57 and older age group (34.4% [CCAHS-1] – 91.4% [CLSA]). Observations for the 18-26-year-old age group were generally low (0.0% [CanPath] – 5.6% [APL]) but were higher for CBS (10.6%) and CCAHS-1 (11.8%). Among studies for which neighborhood deprivation was available, specimen counts across social deprivation quintile were balanced, but only 8.2% (CBS), 8.4% (APL), 9.6% (CLSA), and 13.1% (CCAHS-1) of specimens were provided from the most materially deprived quintile of neighborhoods. Most studies skewed white (78.2% [Ab-C] – 94.7% [CLSA]) and female (52.3% [CLSA] – 65.6% [CanPath]), except CBS which skewed white (81.7%) and male (58.2%). Rural specimens accounted for 8.6% (CanPath) – 17.6% (CCAHS-1) of all specimens across studies. Convenience samples collected substantially more specimens for each demographic strata compared to other recruitment strategies (Supplementary Figures S1-S4).

### Representation ratio analysis

Studies had reasonable representation across sexes (representation ratio 0.7-1.3; Figure 1) and, when available, by social deprivation (Supplementary Figure S5). In all studies, racialized minority subgroups were underrepresented (representation ratio < 1) for some age and sex strata (Figure 2). Racialized minority representation, while still low, was often better in older age groups (Ab-C, CanPath, and CLSA), but was better for younger age groups among women for CBS. Urban regions produced larger representation ratios by age and sex strata than rural regions in all studies (Figures 1-2). While APL had reasonable representation of all material deprivation quintiles and rural residents (representation ratio 0.8-1.3), the CBS population skewed towards less materially deprived neighborhoods and urban regions (though rural representation for CBS was higher than the three longitudinal cohort studies). 18-26-year-old males were underrepresented across most sex and urbanicity strata in all studies for which they were eligible to be sampled.

**Figure 1:**
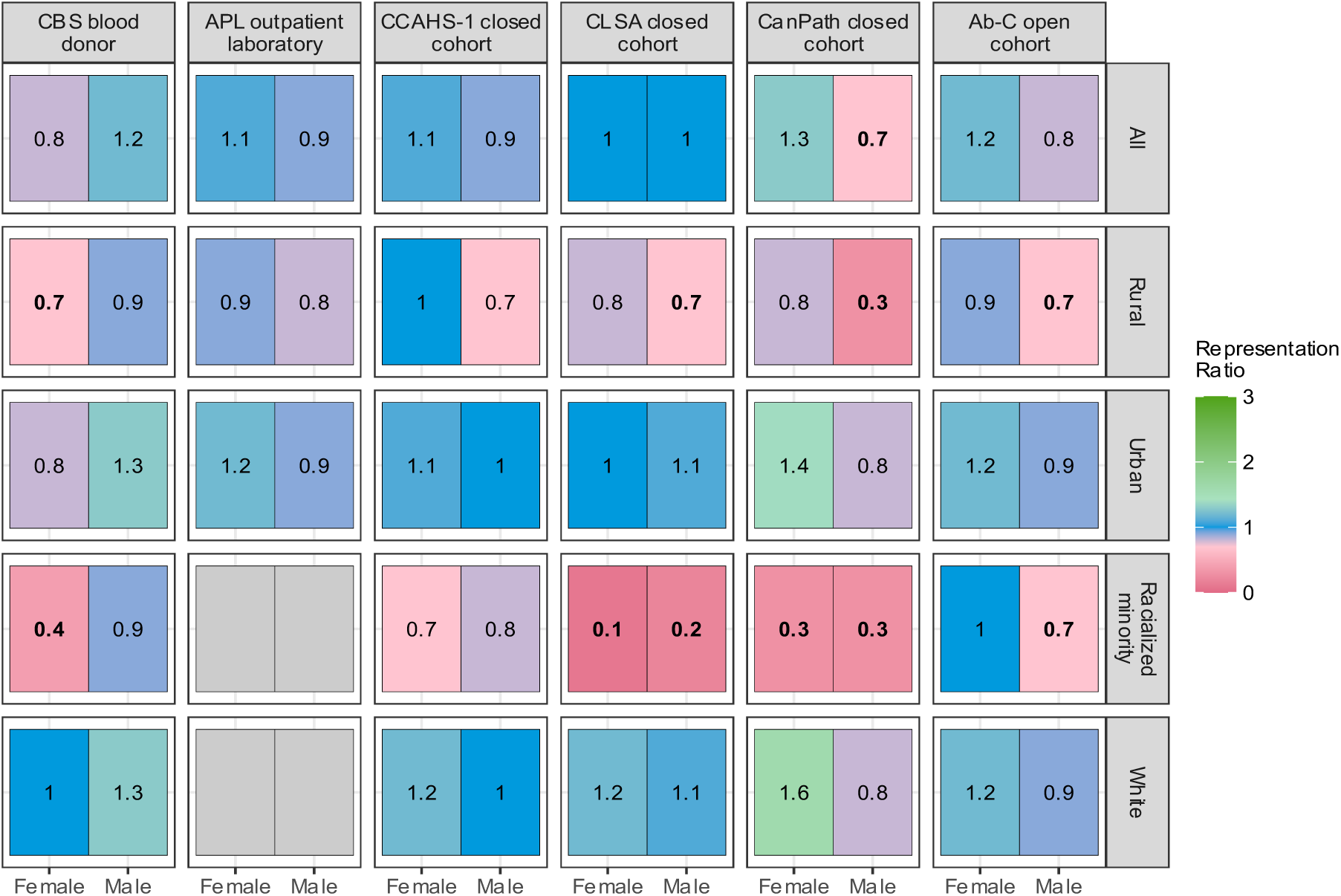
SARS-CoV-2 serological study representativeness by sex, urbanicity, and racial/ethnic identity. Representativeness was calculated by dividing the proportion of study specimens collected from a subgroup by the proportion of general population in the subgroup. Total population counts were estimated using the 2016 Canadian census [19]. Bolded representation ratios indicate greater than 95% of subgroup bootstrap replicates produced representation ratios below 0.75. Bootstrapping was not performed for studies with weighted representation ratios (CCAHS-1).

**Figure 2:**
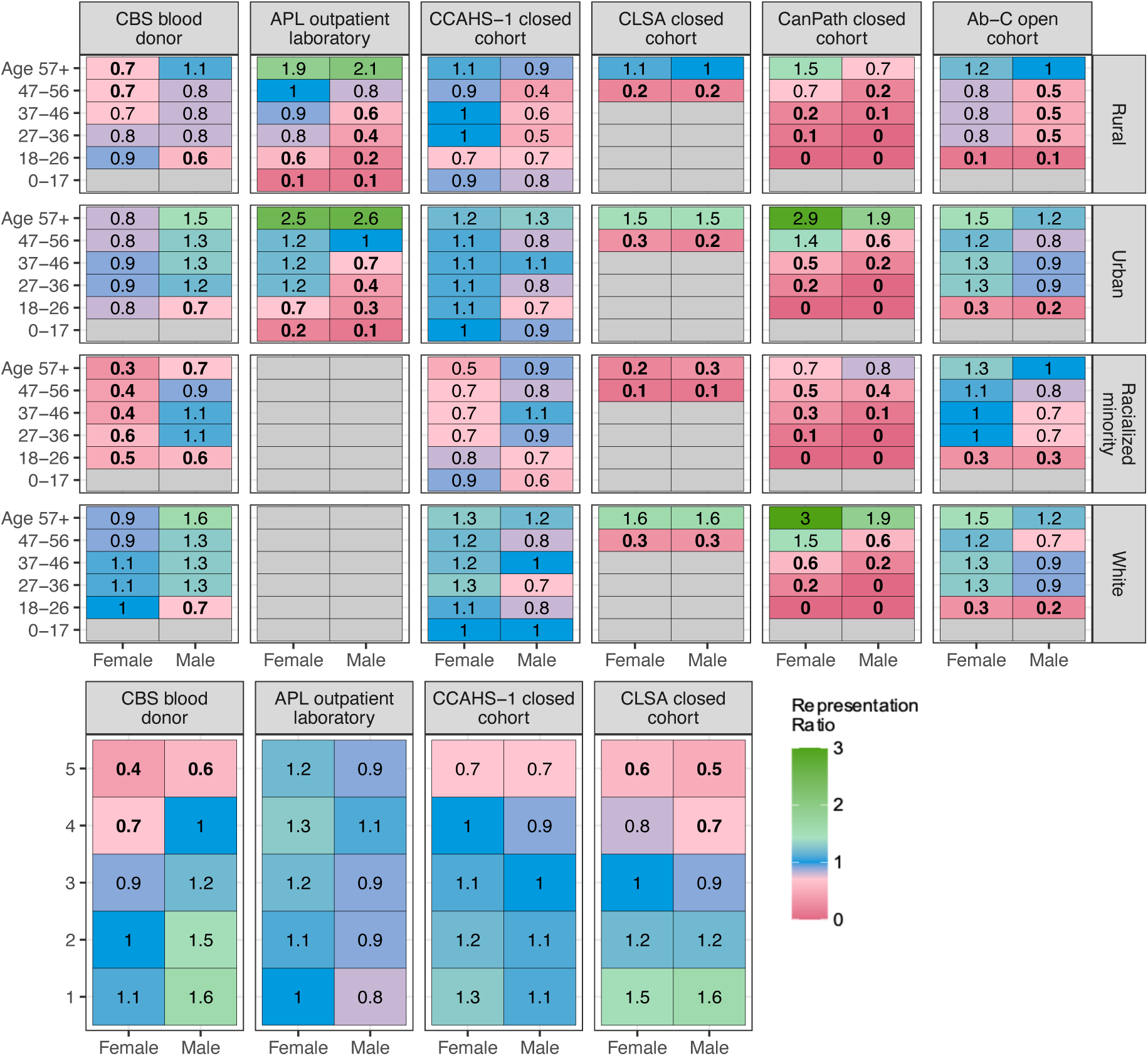
SARS-CoV-2 serological study representativeness by age group, sex, urbanicity, racial/ethnic identity, and material deprivation quintile. Representativeness was calculated by dividing the proportion of study specimens collected from a subgroup by the proportion of general population in the subgroup. Total population counts were estimated using the 2016 Canadian census [19]. Material deprivation scores were not available for Ab-C and CanPath studies. Bolded representation ratios indicate greater than 95% of subgroup bootstrap replicates produced representation ratios below 0.75. Bootstrapping was not performed for studies with weighted counts (CCAHS-1).

Among 18-46-year-olds, specimens from the Ab-C open cohort produced larger representation ratios across sex and urbanicity strata compared to CanPath (Figure 2). Representation ratios of 18-46-year-old rural residents were generally larger across age and sex strata in CCAHS-1 than several studies with probabilistic recruitment strategies (Ab-C, CanPath). Racialized minorities aged 47 years and older were sufficiently represented (representation ratio 0.8-1.3) in the Ab-C open cohort but were underrepresented in the CanPath and CLSA closed cohorts, except for males aged 57 and older in CanPath. Of the two convenience samples, the CBS study was more representative of participants aged 18-46 across sex and urbanicity strata, whereas the APL study was more representative of individuals aged 47 and older. In CCAHS-1, the only study that sampled in the three Canadian territories, 0-17-year-olds were underrepresented across sexes in territorial specimens (Supplementary Figure S6). Bootstrapping revealed low uncertainty in whether a representation ratio was greater or less than one for most subgroups (Supplementary Figures S7-S13). A sensitivity analysis reclassifying mixed race/ethnicity participants as white had little impact except for the Ab-C cohort, for which 55% of racialized minorities were reclassified as white, leading to lower representation ratios (Supplementary Figures S14-S15). Including Indigenous-identifying individuals as racialized minorities in the representation ratio denominator for studies for which data were unavailable (CLSA and CanPath) had little impact on findings (Supplementary Figure S16).

### Sample count analysis

The convenience samples with large overall sample size produced substantially more cells with counts greater than 25 across 4 levels of stratification compared to all other study designs in the primary analysis (Table 2) and sensitivity analysis (Supplementary Table S4). Pre-existing closed probabilistic cohorts (CLSA, CanPath) produced a greater proportion of cells with counts greater than 25 than other probabilistic recruitment strategies (Ab-C, CCAHS-1) for all strata.

**Table 2:**
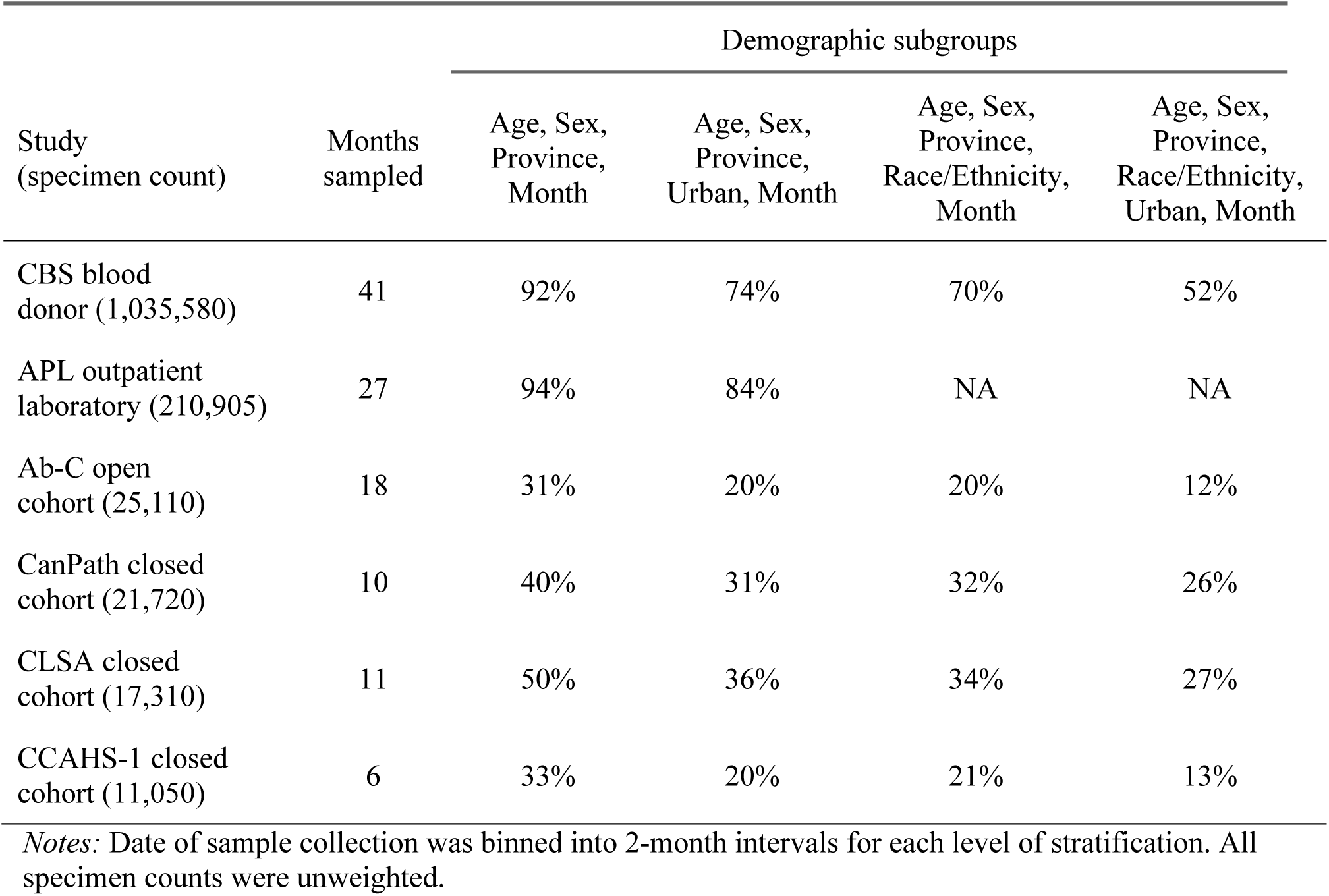
Percentage of SARS-CoV-2 serology study demographic subgroups with greater than 25 collected specimens.

| Study<br>(specimen count) | Months<br>sampled | Demographic subgroups |  |  |  |
| --- | --- | --- | --- | --- | --- |
|  |  | Age, Sex,<br>Province,<br>Month | Age, Sex,<br>Province,<br>Urban, Month | Age, Sex,<br>Province,<br>Race/Ethnicity,<br>Month | Age, Sex,<br>Province,<br>Race/Ethnicity,<br>Urban, Month |
| CBS blood donor (1,035,580) | 41 | 92% | 74% | 70% | 52% |
| APL outpatient laboratory (210,905) | 27 | 94% | 84% | NA | NA |
| Ab-C open cohort (25,110) | 18 | 31% | 20% | 20% | 12% |
| CanPath closed cohort (21,720) | 10 | 40% | 31% | 32% | 26% |
| CLSA closed cohort (17,310) | 11 | 50% | 36% | 34% | 27% |
| CCAHS-1 closed cohort (11,050) | 6 | 33% | 20% | 21% | 13% |

*Notes:* Date of sample collection was binned into 2-month intervals for each level of stratification. All specimen counts were unweighted.

## DISCUSSION

In this study, we developed a simple method for characterizing study population representativeness and applied it to describe the variability in sociodemographic representativeness across six SARS-CoV-2 serosurveillance studies with diverse recruitment strategies. No study was adequately representative of all sociodemographic subgroups.

Representation ratios are a flexible diagnostic measure for characterizing study populations. Ratios can consider any combination of characteristics for which reliable estimates of their distribution in a target population are available. Ratios can be used to compare study populations, even when target populations differ, and can be estimated before and after application of sample weights. Notably, representation ratios do not provide insights into the relationship between participant characteristics and a study’s estimand, and do not address unmeasured confounding within subgroups.

Probabilistic surveys have traditionally been considered the ‘gold standard’ for obtaining representative samples [7]. Use of administrative datasets to construct sampling frames often provides superior population coverage compared to non-probability samples that rely on participant self-selection. The statistical framework also permits estimation of sampling errors and characteristics associated with non-response [22]. While resource constraints may limit the ability of probabilistic designs to perform repeated specimen collection, non-probability sampling within continuous streams of residual blood specimens, such as blood donors, may be more feasible for modelling longitudinal trends, which can also incorporate complex geographic structures [23]. The generalizability of probabilistic designs may also be limited if differences between respondents and non-respondents are non-random [7]. Bias may be introduced via a ‘healthy volunteer’ effect whereby cohort participants are healthier than the general population [24], similar to the ‘healthy donor’ bias documented in blood donor research cohorts [25]. Where available, response rates of the included studies were fairly low (23% [CCAHS-1], 25% [Ab-C] [6,12]; these response rates exclude individuals who completed a questionnaire but did not provide a blood sample). This suggests non-response bias could partially explain some of the observed differences in representativeness between study designs. The above response rates are consistent with other probabilistic serosurveys [11], although response rates as high as 69% have been reported [10].

Many large-scale SARS-CoV-2 studies relied on blood donor and healthcare patient populations for serology specimens [22,26]. Blood donors have been discounted as a population for public health surveillance, while the potential for expanded screening of residual outpatient laboratory samples remains unclear. However, we found the representativeness of these convenience populations compared favorably to other designs for some sociodemographic dimensions. For low- and middle-income countries with limited operational resources, leveraging residual blood samples may provide a cost-effective avenue to obtain representative data. Future studies should evaluate the potential of linkage to administrative datasets to better characterize representativeness and derive statistical weights for adjustment. Gaps in demographic representation may be overcome by synthesizing data from multiple surveillance streams, though differences in choice of assay, use of venous blood draws or dried blood samples, and the format or availability of variables can curtail the ability to synthesize data across studies [27,28].

Racialized minorities were underrepresented across all studies. Language barriers and skepticism of research or medical institutions may contribute to poor representation of some minority groups [29,30]. While use of stratified random sampling or sampling weights may improve sample representativeness, they do not address the underlying individual and societal factors governing participation in health research. Direct engagement and collaboration with community members throughout the research cycle may mitigate recruitment barriers by facilitating trust, reducing misinformation, and ensuring study materials are accessible [30,31]. Racialized minorities may be better represented in healthcare cohorts like APL [32], though a lack of race-based data in Canadian administrative healthcare datasets may make this difficult to measure [33]. Notably, representation of racialized minorities improved as age increased in most studies requiring participant opt-in (Ab-C, CLSA, CanPath), but young minorities exhibited better representativeness compared to older subgroups in CBS. Lack of a standardized definition of participant race/ethnicity impeded comparison across studies and prevented assessment of representativeness by specific minority group.

Several other dimensions of representativeness varied across studies. The Ab-C open cohort was substantially more representative of 18-46-year-olds across sex and urbanicity strata compared to the CanPath longitudinal closed cohort (Figure 2). CLSA and CanPath recruited participants aged 45-85 in 2010 and 35-74 in 2009, respectively, leading to older age distributions for their COVID-19 sub-studies [15,16]. Between convenience samples, individuals residing in highly materially deprived areas were underrepresented when using blood donations (representation ratio 0.4–0.6), but not when using outpatient labs (representation ratio 0.9-1.2). Donor eligibility criteria, along with the ‘healthy donor effect’ or other unmeasured socioeconomic factors, may homogenize the demographic composition of the sampled donor pool [25]. Rural regions were consistently underrepresented compared to urban counterparts in all studies, which may be due to urban-centric recruitment patterns or willingness to travel for specimen collection (Figures 1-2).

The study had several limitations. First, our analysis considered representativeness by age, sex, race/ethnicity, urbanicity, and neighborhood deprivation. Many other sociodemographic dimensions are important considerations for representativeness in serosurveillance studies, particularly those related to health and disability. We hypothesize a ‘healthy participant’ sampling bias may have led to underrepresentation of individuals with poor health and/or disability in all study populations except outpatient laboratories [24,25]. Prior analyses of the pre-existing longitudinal cohorts included in our study indicated participants are more educated and/or have higher income than the general population [12,15,16], as are blood donors in the United States [34]. Second, the measurement of race/ethnicity differed between studies.

Race/ethnicity options for CBS included four mutually exclusive categories, while the Ab-C, CCAHS-1, CanPath, CLSA, and census datasets permitted selection of multiple racial/ethnic identities. This necessitated dichotomizing the race/ethnicity variable as white or racialized minority and may have biased the CBS representation estimate if individuals who identified as mixed race/ethnicity selected their race/ethnicity as white during donation. Additionally, due to unavailable Indigenous identity data, we modified our representation assessment for the CLSA and CanPath studies by omitting Indigenous-identifying individuals from the census dataset. Our sensitivity analysis suggests this did not substantially impact our findings (Supplementary Figure S16). Third, we restricted our analysis to SARS-CoV-2 serostudies conducted within a single country and used an acceptability level of underrepresentation that is context-specific and open to interpretation. We focused on strata with fewer than 25 samples in our cell count analysis, but this threshold was largely arbitrary. Fourth, we did not analyze factors shaping the sociodemographic composition of each study, including intentional oversampling. For example, CCAHS-1 used a stratified random sampling strategy that oversampled geographic regions with greater COVID-19 prevalence and less populated regions to improve estimate precision. Less populated areas of Canada often have fewer racialized minorities, which likely contributes to lower representation ratios [6]. Understanding the causes and consequences of each study’s sociodemographic composition requires more detailed analysis than is presented here. Finally, our study is not a comprehensive assessment of all SARS-CoV-2 serology studies conducted in Canada. Demographic groups excluded here were evaluated elsewhere [35].

Understanding variability in demographic representation between study designs is an important consideration when planning serosurveillance studies, which increasingly leverage pre-existing samples or cohorts. This study provides a simple metric to evaluate and compare the representativeness of study populations. We found that underrepresentation of racialized minorities and younger age groups was common and not restricted to convenience samples, which had better representation for some sociodemographic strata. This suggests that representative estimates could be obtained in resource-constrained settings by leveraging lower-cost approaches, such as existing blood or laboratory services, compared to large-scale probabilistically sampled serosurveys. Identifying coverage barriers is vital to support adequate representation and detection of disease trends within demographic subgroups. We also observed differences in the measurement of participant race/ethnicity between studies. This highlights the necessity for consistent, and sufficient, measurement of sociodemographic variables, along with the need to adopt a standardized approach to the measurement of self-identified race/ethnicity in Canada.

## Supporting information

STROBE checklist

## Data Availability

The authors are not authorized to share individual-level data from any study. Processes are available for researchers to request access to datasets for studies that have undergone institutional ethical approval. Data from Canadian Blood Services and Alberta Precision Laboratories may be made available upon request, subject to internal review, privacy legislation, data sharing agreements, and research ethics approval. The CCAHS-1 study by Statistics Canada can be analyzed for approved projects at Research Data Centres located across Canada (https://www.statcan.gc.ca/en/microdata/data-centres/access). Data are available from the Canadian Longitudinal Study on Aging (www.clsa-elcv.ca) for researchers who meet the criteria for access to de-identified CLSA data. Access to the Ab-C study data can be requested through the COVID-19 Immunity Task Force Databank (https://portal.citf.mcgill.ca/). Access to the CanPath data can be requested through the CanPath data portal (https://portal.canpath.ca/). Analytical code will be available in a public repository upon publication.

## ABBREVIATIONS

Ab-C: Action to Beat Coronavirus
APL: Alberta Precision Laboratories
CanPath: Canadian Partnership for Tomorrow’s Health
CBS: Canadian Blood Services
CCAHS-1: Canadian COVID-19 Antibody and Health Survey 1
CLSA: Canadian Longitudinal Study on Aging
SARS-CoV-2: Severe acute respiratory syndrome coronavirus-2

## DECLARATIONS

### Ethics approval and consent to participate

All studies analyzed were approved by a Research Ethics Board or Institutional Review Board of a Canadian institution as reported previously. This secondary analysis of six studies was approved by the McGill University Faculty of Medicine and Health Sciences Research Ethics Board (study number 22-03-077).

### Consent for participation

Not applicable.

### Authors’ contributions

MJK and WAR designed the study with input from DLB, SFO, and CC. MJK, YY, and JC contributed to data analysis. MJK and WAR drafted the initial manuscript. All authors revised the manuscript and approved the final version.

## Acknowledgements

This research was made possible using data collected by Canadian Blood Services’ SARS-CoV-2 Seroprevalence Study; the Canadian Partnership for Tomorrow’s Health (CanPath – formerly CPTP) COVID-19 Antibody study and its regional cohorts the BC Generations Project, Alberta’s Tomorrow Project, the Ontario Health Study, CARTaGENE, Manitoba Tomorrow Project and the Atlantic Partnership for Tomorrow’s Health; the Action to Beat Coronavirus in Canada (Ab-C) study; the Alberta Precision Laboratories study; the Canadian COVID-19 Antibody and Health Survey (CCAHS) conducted by Statistics Canada, and the Canadian Longitudinal Study on Aging (CLSA). We thank the participants, staff, and researchers of all six studies. We also thank Scott McLeish for the insightful discussions which greatly improved the quality of the manuscript. Funding for the CLSA is provided by the Government of Canada through the Canadian Institutes of Health Research (CIHR) under grant reference: LSA 94473 and the Canada Foundation for Innovation, as well as the provinces, Newfoundland, Nova Scotia, Quebec, Ontario, Manitoba, Alberta, and British Columbia. Funding for support of the CLSA COVID-19 questionnaire-based study is provided by the Juravinski Research Institute, Faculty of Health Sciences, McMaster University, the Provost Fund from McMaster University, the McMaster Institute for Research on Aging, the Public Health Agency of Canada/CIHR grant reference CMO 174125 and the government of Nova Scotia. This research has been conducted using the CLSA Baseline Comprehensive Dataset v7.0, Baseline Tracking Dataset v4.0, Follow-up 1 Comprehensive Dataset v4.0, Follow-up 1 Tracking Dataset v3.0, Follow-up 2 Comprehensive Dataset v1.0, Follow-up 2 Tracking Dataset v1.0, COVID-19 Questionnaire Study Dataset v1.1, and COVID-19 Seroprevalence Study Dataset v1.0 under Application Number 2209005. The CLSA is led by Drs. Parminder Raina, Christina Wolfson, and Susan Kirkland. We appreciate the funding that made each study possible, including the support each study received from the Government of Canada through the COVID-19 Immunity Task Force. The opinions expressed in this manuscript are the author’s own and do not reflect the views of the data providers and/or their institutions.

## REFERENCES

1. O’Brien SF, Caffrey N, Yi QL, et al. Cross-Canada Variability in Blood Donor SARS-CoV-2 Seroprevalence by Social Determinants of Health. Microbiology Spectrum. 2023;11(1):e03356–22. 10.1128/spectrum.03356-22

2. Charlton CL, Nguyen LT, Bailey A, et al. Pre-Vaccine Positivity of SARS-CoV-2 Antibodies in Alberta, Canada during the First Two Waves of the COVID-19 Pandemic. Microbiology Spectrum. 2021;9(1):e00291–21. 10.1128/Spectrum.00291-21

3. Jha, Prabhat. Action to beat Coronavirus study: Dataset contributed to the CITF Databank. Version DRAFT. Borealis. doi:10.5683/SP3/LA2IKO

4. Canadian Longitudinal Study on Aging. Data Support Document SARS-CoV-2 Antibodies. 2024.

5. Canadian Partnership for Tomorrow’s Health. COVID-19 initiatives. CanPath - Canadian Partnership for Tomorrow’s Health. Available from: https://canpath.ca/covid-19-initiatives/.

6. Statistics Canada. Canadian COVID-19 Antibody and Health Survey (CCAHS). 2020. Available from: https://www23.statcan.gc.ca/imdb/p2SV.pl?Function=getSurvey&Id=1287991

7. Cornesse C, Bosnjak M. Is there an association between survey characteristics and representativeness? A meta-analysis. Survey Research Methods. 2018;12(1):1–13. 10.18148/srm/2018.v12i1.7205

8. Offergeld R, Preußel K, Zeiler T, et al. Monitoring the SARS-CoV-2 Pandemic: Prevalence of Antibodies in a Large, Repetitive Cross-Sectional Study of Blood Donors in Germany—Results from the SeBluCo Study 2020–2022. Pathogens. 2023;12(4):551. 10.3390/pathogens12040551

9. Bogogiannidou Z, Vontas A, Dadouli K, et al. Repeated leftover serosurvey of SARS-CoV-2 IgG antibodies, Greece, March and April 2020. Eurosurveillance. 2020;25(31):2001369. 10.2807/1560-7917.ES.2020.25.31.2001369

10. Pérez-Gómez B, Pastor-Barriuso R, Fernández-de-Larrea N, et al. SARS-CoV-2 Infection During the First and Second Pandemic Waves in Spain: The ENE–COVID Study. Am J Public Health. 2023;113(5):533–544. 10.2105/AJPH.2023.307233

11. Ward H, Atchison C, Whitaker M, et al. Design and Implementation of a National Program to Monitor the Prevalence of SARS-CoV-2 IgG Antibodies in England Using Self-Testing: The REACT-2 Study. Am J Public Health. 2023;113(11):1201–1209. 10.2105/AJPH.2023.307381

12. Tang X, Sharma A, Pasic M, et al. Assessment of SARS-CoV-2 Seropositivity During the First and Second Viral Waves in 2020 and 2021 Among Canadian Adults. JAMA Network Open. 2022;5(2):e2146798. 10.1001/jamanetworkopen.2021.46798

13. Patel EU, Bloch EM, Tobian AAR. Severe Acute Respiratory Syndrome Coronavirus 2 Serosurveillance in Blood Donor Populations. The Journal of Infectious Diseases. 2022;225(1):1–4. 10.1093/infdis/jiab517

14. Rudolph JE, Zhong Y, Duggal P, et al. Defining representativeness of study samples in medical and population health research. BMJ Med. 2023;2(1):e000399. 10.1136/bmjmed-2022-000399

15. Raina P, Wolfson C, Kirkland S, et al. Cohort Profile: The Canadian Longitudinal Study on Aging (CLSA). International Journal of Epidemiology. 2019;48(6):1752–1753j. 10.1093/ije/dyz173

16. Dummer TJB, Awadalla P, Boileau C, et al. The Canadian Partnership for Tomorrow Project: A pan-Canadian platform for research on chronic disease prevention. CMAJ. 2018;190(23):E710–E717. 10.1503/cmaj.170292

17. von Elm E, Altman DG, Egger M, et al. The Strengthening the Reporting of Observational Studies in Epidemiology (STROBE) Statement: Guidelines for reporting observational studies. Epidemiology. 2007;18(6):800–804.

18. Pampalon R, Hamel D, Gamache P, et al. An area-based material and social deprivation index for public health in Québec and Canada. Can J Public Health. 2012;103(8 Suppl 2):S17–22. 10.1007/BF03403824

19. Statistics Canada. Census Profile, 2016 Census. 2017. Available from: https://www12.statcan.gc.ca/census-recensement/2016/dp-pd/prof/index.cfm?Lang=E

20. Downes M, Carlin JB. Multilevel regression and poststratification as a modeling approach for estimating population quantities in large population health studies: A simulation study. Biometrical Journal. 2020;62(2):479–491. 10.1002/bimj.201900023

21. R Core Team. R: A Language and Environment for Statistical Computing. Vienna, Austria: R Foundation for Statistical Computing; 2023. https://www.R-project.org/.

22. Lindan CP, Desai M, Boothroyd D, et al. Design of a population-based longitudinal cohort study of SARS-CoV-2 incidence and prevalence among adults in the San Francisco Bay Area. Annals of Epidemiology. 2022;67:81–100. 10.1016/j.annepidem.2021.11.001

23. Yu Y, Knight MJ, Gibson D, et al. Temporal trends in disparities in COVID-19 seropositivity among Canadian blood donors. International Journal of Epidemiology. 2024;53(3):dyae078. 10.1093/ije/dyae078

24. Froom P, Melamed S, Kristal-Boneh E, et al. Healthy Volunteer Effect in Industrial Workers. Journal of Clinical Epidemiology. 1999;52(8):731–735. 10.1016/S0895-4356(99)00070-0

25. Atsma F, Veldhuizen I, Verbeek A, et al. Healthy donor effect: Its magnitude in health research among blood donors. Transfusion. 2011;51(8):1820–1828. doi:10.1111/j.1537-2995.2010.03055.x

26. Anand S, Montez-Rath M, Han J, et al. Prevalence of SARS-CoV-2 antibodies in a large nationwide sample of patients on dialysis in the USA: A cross-sectional study. The Lancet. 2020;396(10259):1335–1344. doi:10.1016/S0140-6736(20)32009-2

27. Mulchandani R, Brown B, Brooks T, et al. Use of dried blood spot samples for SARS-CoV-2 antibody detection using the Roche Elecsys ® high throughput immunoassay. Journal of Clinical Virology. 2021;136:104739. 10.1016/j.jcv.2021.104739

28. Patel EU, Bloch EM, Clarke W, et al. Comparative Performance of Five Commercially Available Serologic Assays To Detect Antibodies to SARS-CoV-2 and Identify Individuals with High Neutralizing Titers. Journal of Clinical Microbiology. 2021;59(2):10.1128/jcm.02257-20. 10.1128/jcm.02257-20

29. Etti M, Fofie H, Razai M, et al. Ethnic minority and migrant underrepresentation in Covid-19 research: Causes and solutions. eClinicalMedicine. 2021;36. 10.1016/j.eclinm.2021.100903

30. Bonevski B, Randell M, Paul C, et al. Reaching the hard-to-reach: A systematic review of strategies for improving health and medical research with socially disadvantaged groups. BMC Medical Research Methodology. 2014;14(1):42. 10.1186/1471-2288-14-42

31. Ekezie W, Czyznikowska BM, Rohit S, et al. The views of ethnic minority and vulnerable communities towards participation in COVID-19 vaccine trials. Journal of Public Health. 2021;43(2):e258–e260. 10.1093/pubmed/fdaa196

32. Anand S, Montez-Rath M, Han J, et al. Prevalence of SARS-CoV-2 antibodies in a large nationwide sample of patients on dialysis in the USA: A cross-sectional study. The Lancet. 2020;396(10259):1335–1344. 10.1016/S0140-6736(20)32009-2

33. Khan MM, Kobayashi K, Vang ZM, et al. Are visible minorities “invisible” in Canadian health data and research? A scoping review. International Journal of Migration, Health and Social Care. 2017;13(1):126–143. 10.1108/IJMHSC-10-2015-0036

34. Patel EU, Bloch EM, Grabowski MK, et al. Sociodemographic and behavioral characteristics associated with blood donation in the United States: A population-based study. Transfusion. 2019;59(9):2899–2907. 10.1111/trf.15415

35. Atkinson A, Albert A, McClymont E, et al. Canadian SARS-CoV-2 serological survey using antenatal serum samples: A retrospective seroprevalence study. Canadian Medical Association Open Access Journal. 2023;11(2):E305–E313. 10.9778/cmajo.20220045

