## Supplemental material for "Sociodemographic characteristics of SARS-CoV-2 serosurveillance studies with diverse recruitment strategies, Canada, 2020 to 2023"

**SUPPLEMENTAL FIGURES**

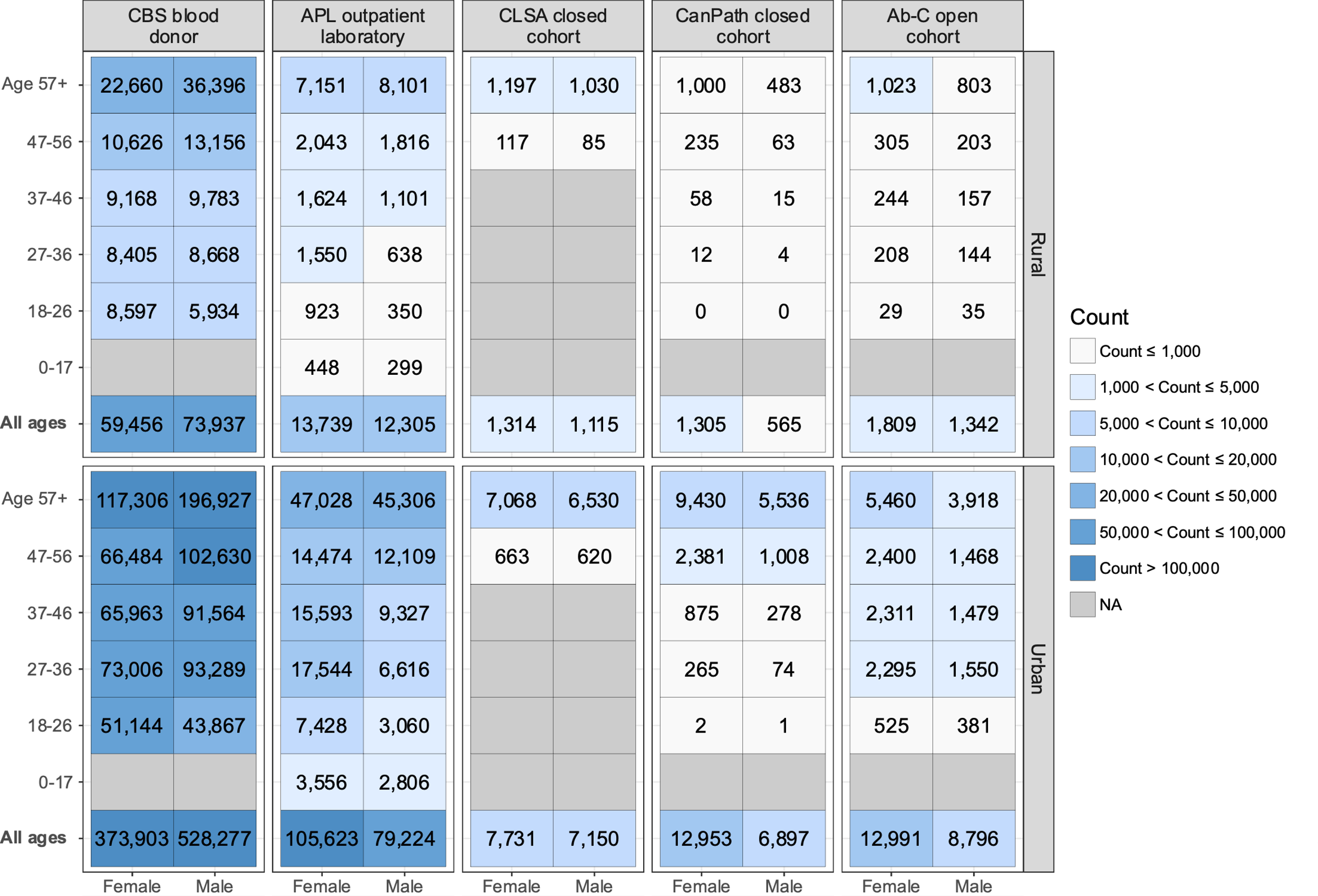

**Supplementary Figure S1: Demographic composition of Canadian SARS-CoV-2 serological studies by age, sex, and urbanicity.** Counts were calculated as the number of serological specimens contributed by each study subgroup. CBS tested 3410 donations from 17-year-old donors, but they were excluded from our analysis. CCAHS-1 counts were not included due to privacy regulations.

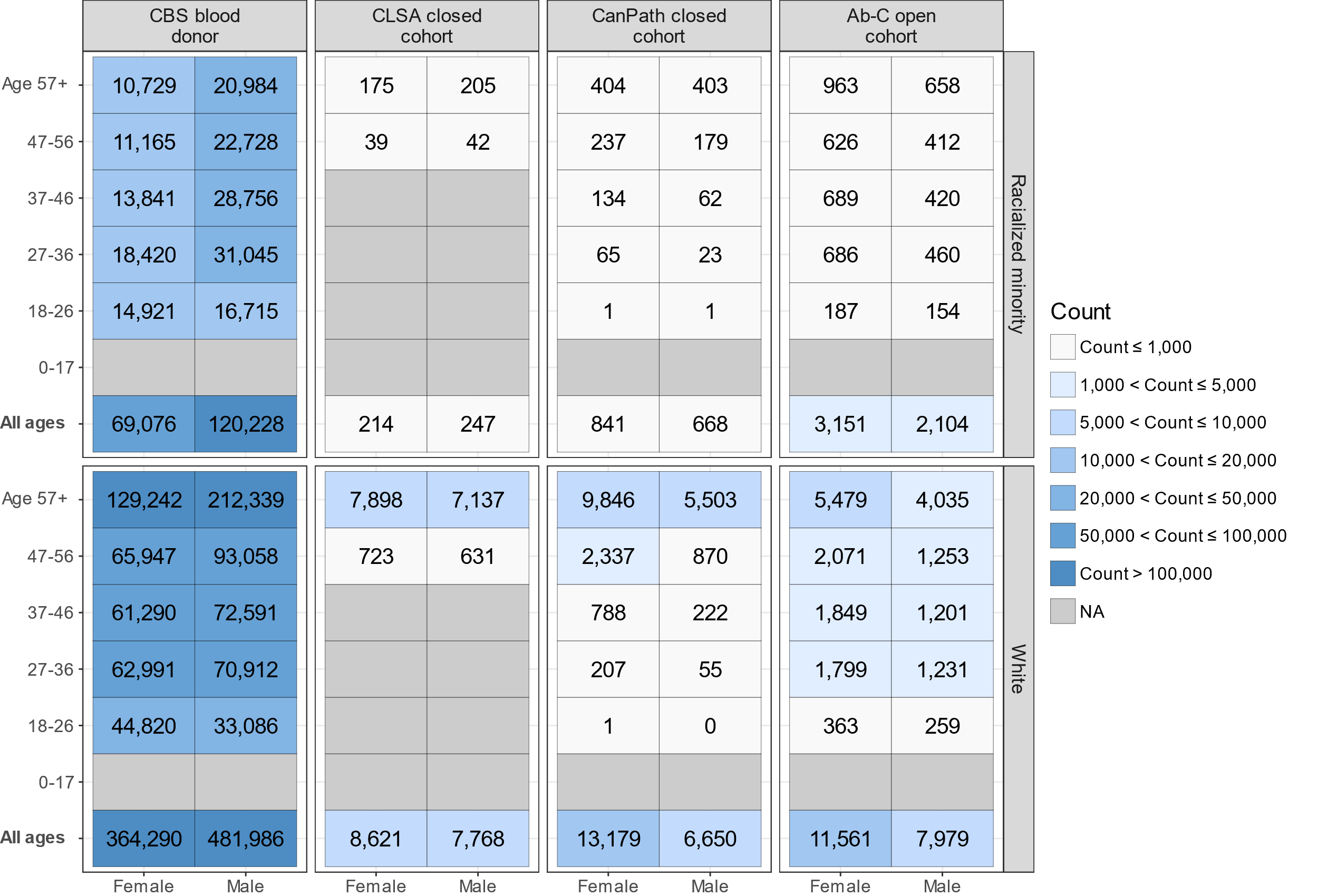

**Supplementary Figure S2: Demographic composition of Canadian SARS-CoV-2 serological studies by age, sex, and self-identified race/ethnicity.** Counts were calculated as the number of serological specimens contributed by each study subgroup. CBS tested 3410 donations from 17-year-old donors, but they were excluded from our analysis. CCAHS-1 counts were not included due to privacy regulations.

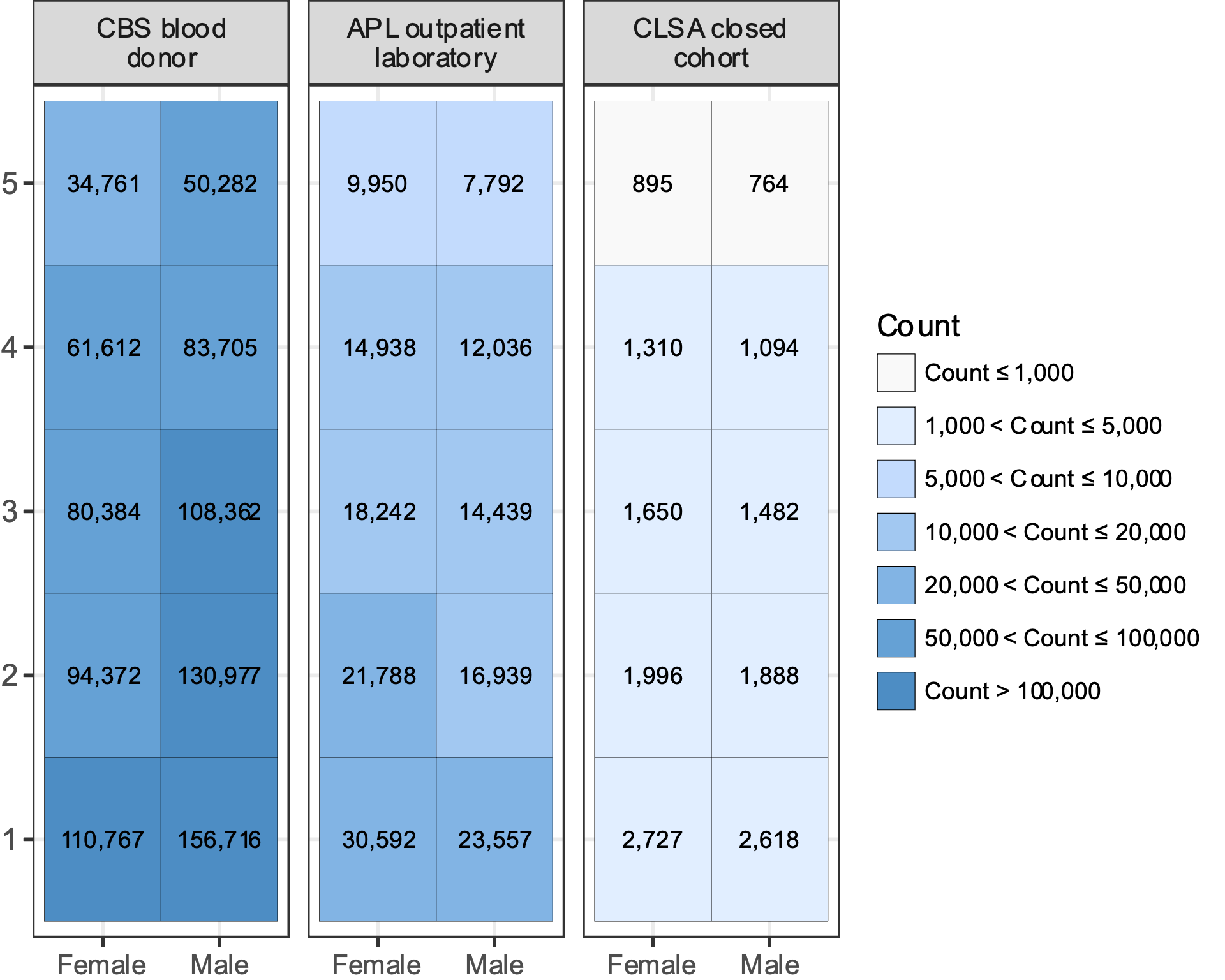

**Supplementary Figure S3: Sociodemographic composition of Canadian SARS-CoV-2 serological studies by sex and material deprivation quintile score.** Counts were calculated as the number of serological specimens contributed by each study subgroup. Scores of 1 and 5 indicate the lowest and highest quintiles of deprivation, respectively. CCAHS-1 counts were not included due to privacy regulations.

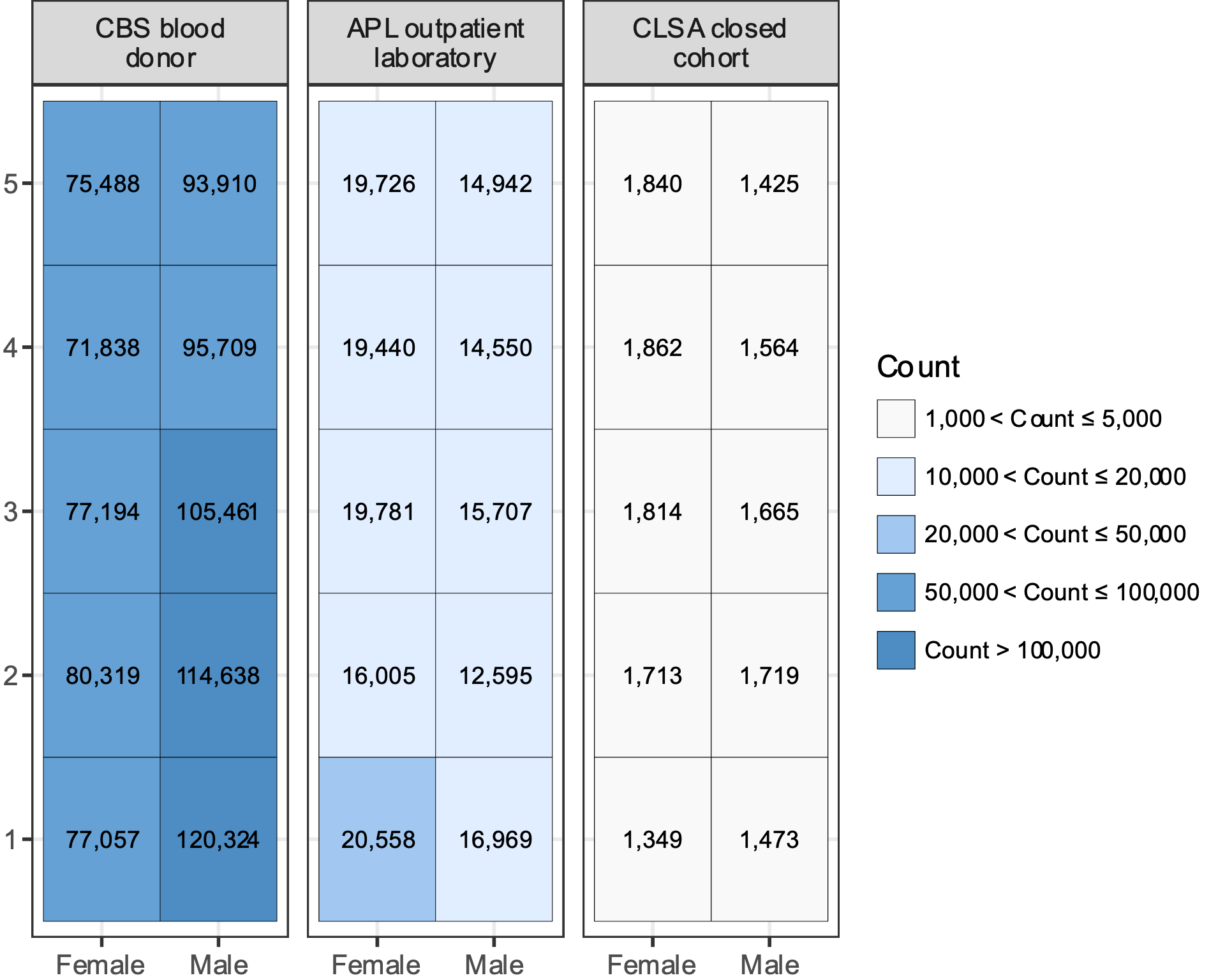

**Supplementary Figure S4: Sociodemographic composition of Canadian SARS-CoV-2 serological studies by sex and social deprivation quintile score.** Counts were calculated as the number of serological specimens contributed by each study subgroup. Scores of 1 and 5 indicate the lowest and highest quintiles of deprivation, respectively. CCAHS-1 counts were not included due to privacy regulations.

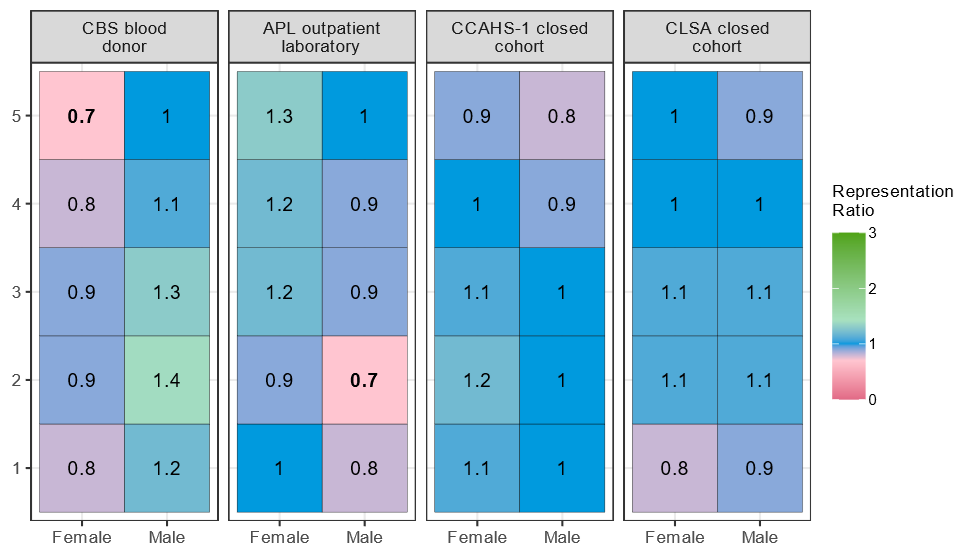

**Supplementary Figure S5: SARS-CoV-2 serological study representativeness by sex and social deprivation quintile.** Representativeness was calculated by dividing the proportion of study specimens collected from a subgroup by the proportion of general population in the subgroup. Total population counts were estimated using the 2016 Canadian census [19]. Social deprivation scores were not available for Ab-C and CanPath studies. Bolded representation ratios indicate greater than 95% of subgroup bootstrap replicates produced representation ratios below 0.75. Bootstrapping was not performed for studies with weighted counts (CCAHS-1).

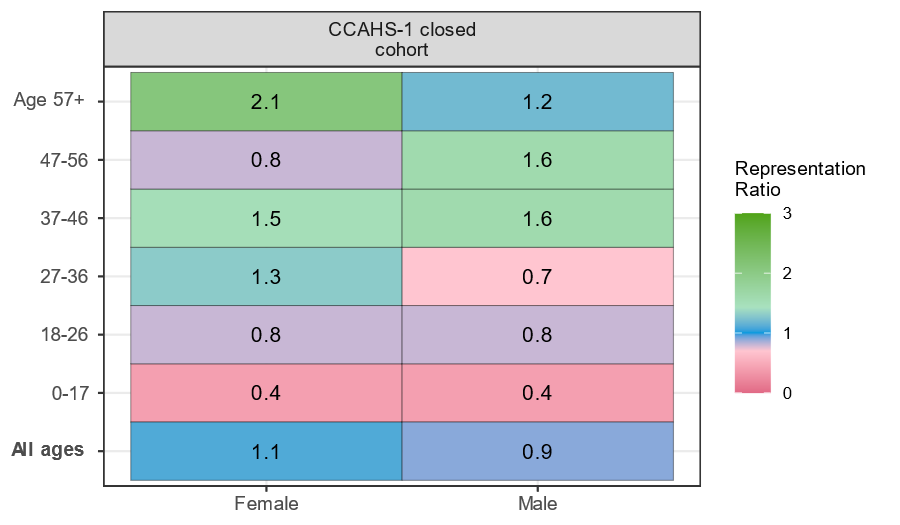

**Supplementary Figure S6: Representativeness of CCAHS-1 study specimens collected from Canadian territories by age group and sex.** Serological specimens were collected from the capital cities of the Canadian territories. Representativeness was calculated by dividing the proportion of study specimens collected from a subgroup by the proportion of population in the subgroup. Total population counts were estimated using the 2016 Canadian census [19]. Bootstrapping was not performed because representation ratios were calculated using weighted study counts. The population distribution of the weighted CCAHS-1 data was assumed to reflect the total territorial population distribution for this analysis.

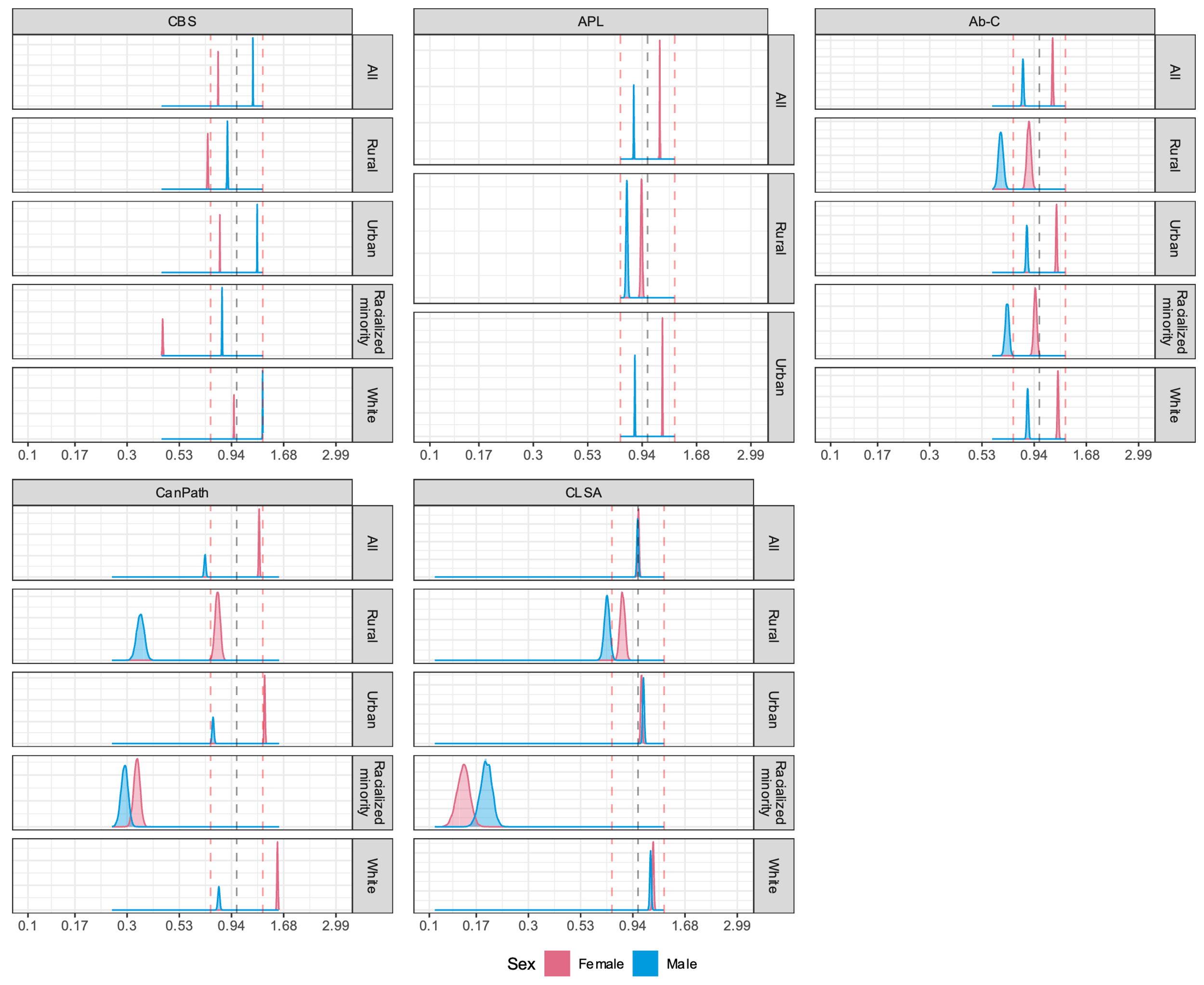

**Supplementary Figure S7: Bootstrap distribution of log-10 transformed representation ratios stratified by sex, urbanicity, and racial/ethnic identity.** Distributions were generated using 5000 bootstrap resamples and plotted with a binwidth of 0.01. Representativeness was calculated by dividing the proportion of study specimens collected from a subgroup by the proportion of population in the subgroup. Total population counts were estimated using the 2016 Canadian census [19]. Dashed red lines indicate representation ratio values of 0.75 and 1.33.

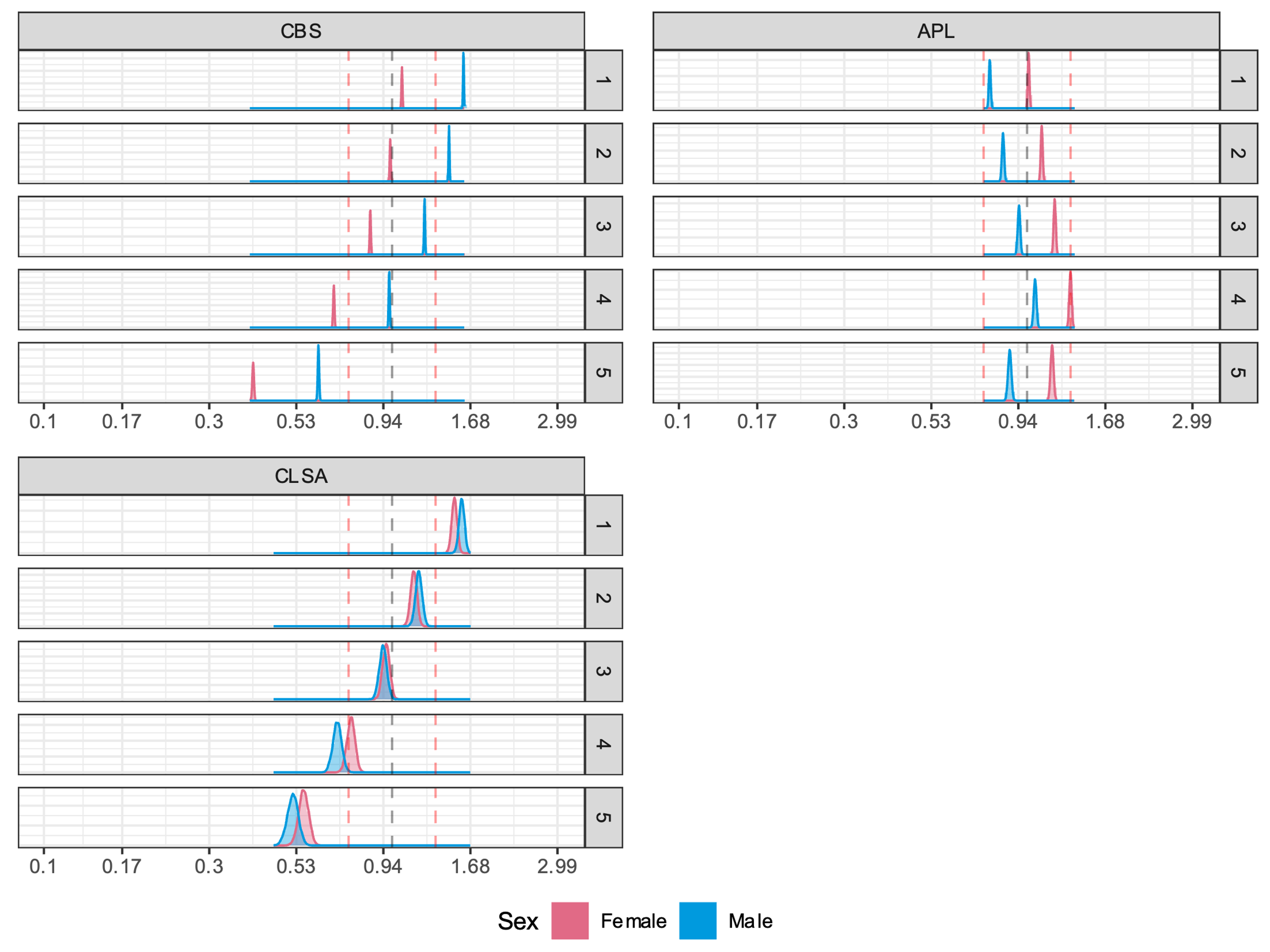

**Supplementary Figure S8: Bootstrap distribution of log-10 transformed representation ratios stratified by sex and material deprivation quintile.** Distributions were generated using 5000 bootstrap resamples and plotted with a binwidth of 0.01. Representativeness was calculated by dividing the proportion of study specimens collected from a subgroup by the proportion of population in the subgroup. Total population counts were estimated using the 2016 Canadian census [19]. Dashed red lines indicate representation ratio values of 0.75 and 1.33.

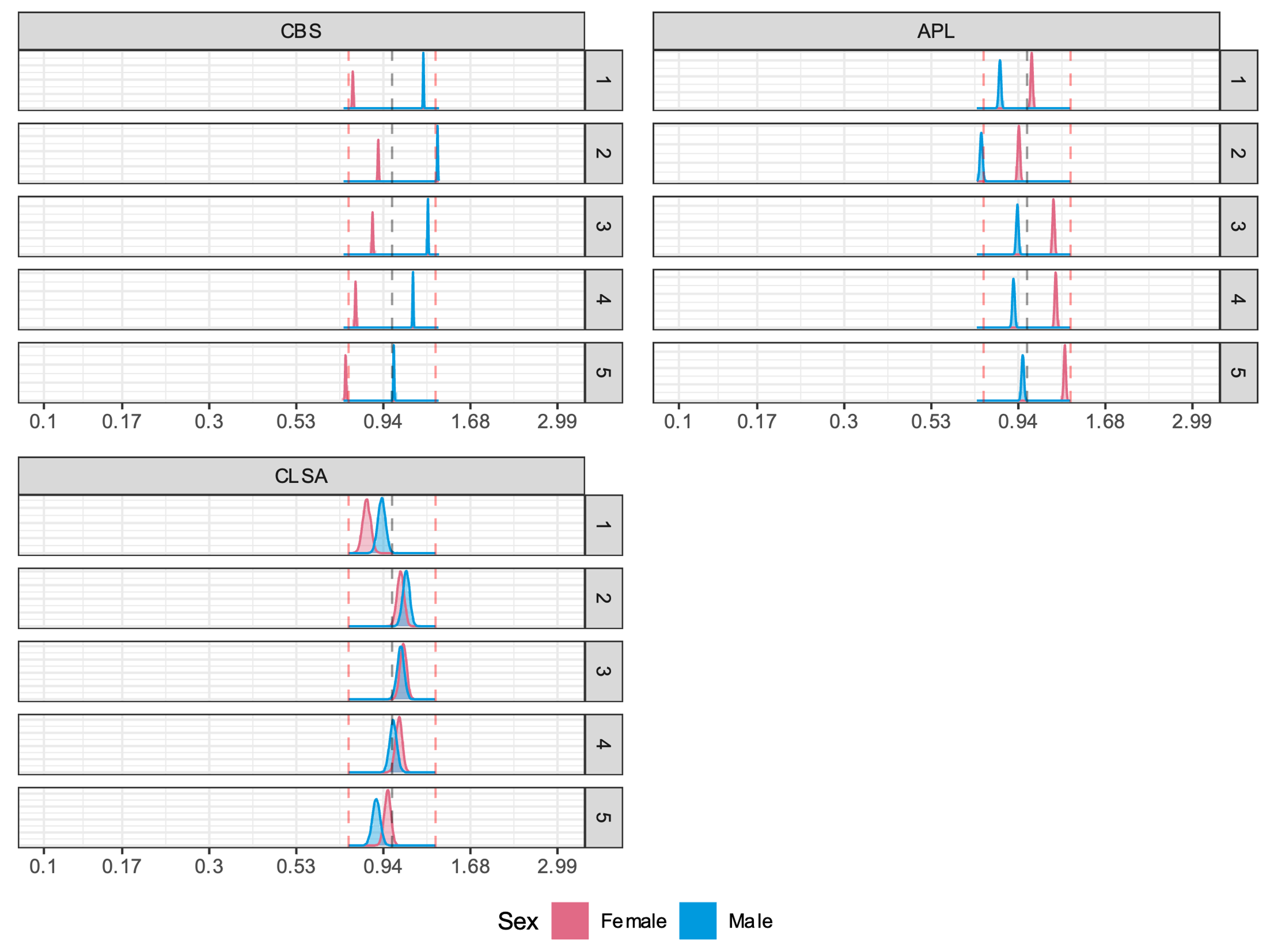

**Supplementary Figure S9: Bootstrap distribution of log10-transformed representation ratios stratified by sex and social deprivation quintile.** Distributions were generated using 5000 bootstrap resamples and plotted with a binwidth of 0.01. Representativeness was calculated by dividing the proportion of study specimens collected from a subgroup by the proportion of population in the subgroup. Total population counts were estimated using the 2016 Canadian census [19]. Dashed red lines indicate representation ratio values of 0.75 and 1.33.

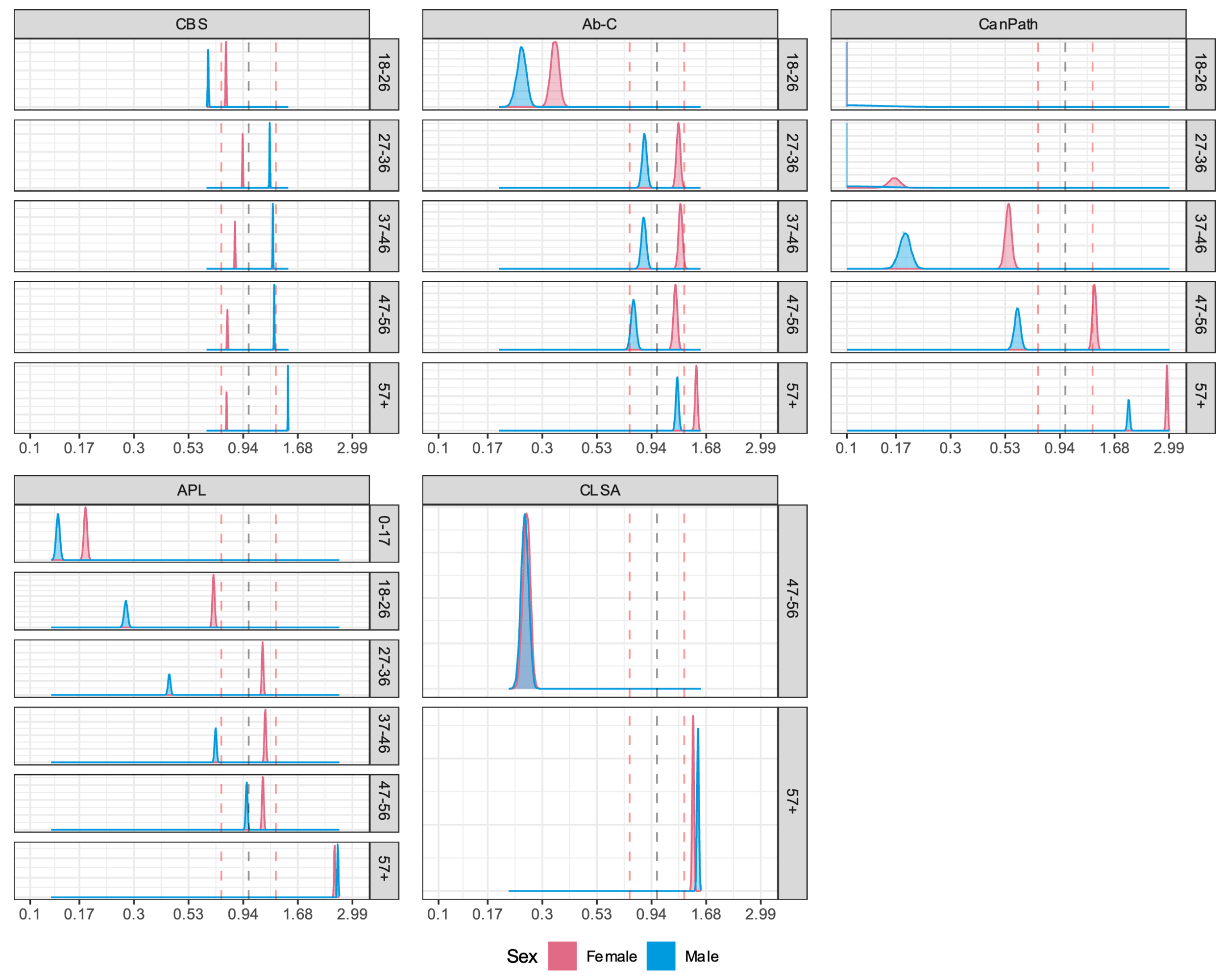

**Supplementary Figure S10: Bootstrap distribution of log10-transformed representation ratios stratified by age group, sex, and urban residence.** Distributions were generated using 5000 bootstrap resamples and plotted with a binwidth of 0.01. Representation ratios between 0.00 – 0.09 were pseudo-adjusted to a value of 0.1 prior to transformation for visualization purposes. Representativeness was calculated by dividing the proportion of study specimens collected from a subgroup by the proportion of population in the subgroup. Total population counts were estimated using the 2016 Canadian census [19]. Dashed red lines indicate representation ratio values of 0.75 and 1.33.

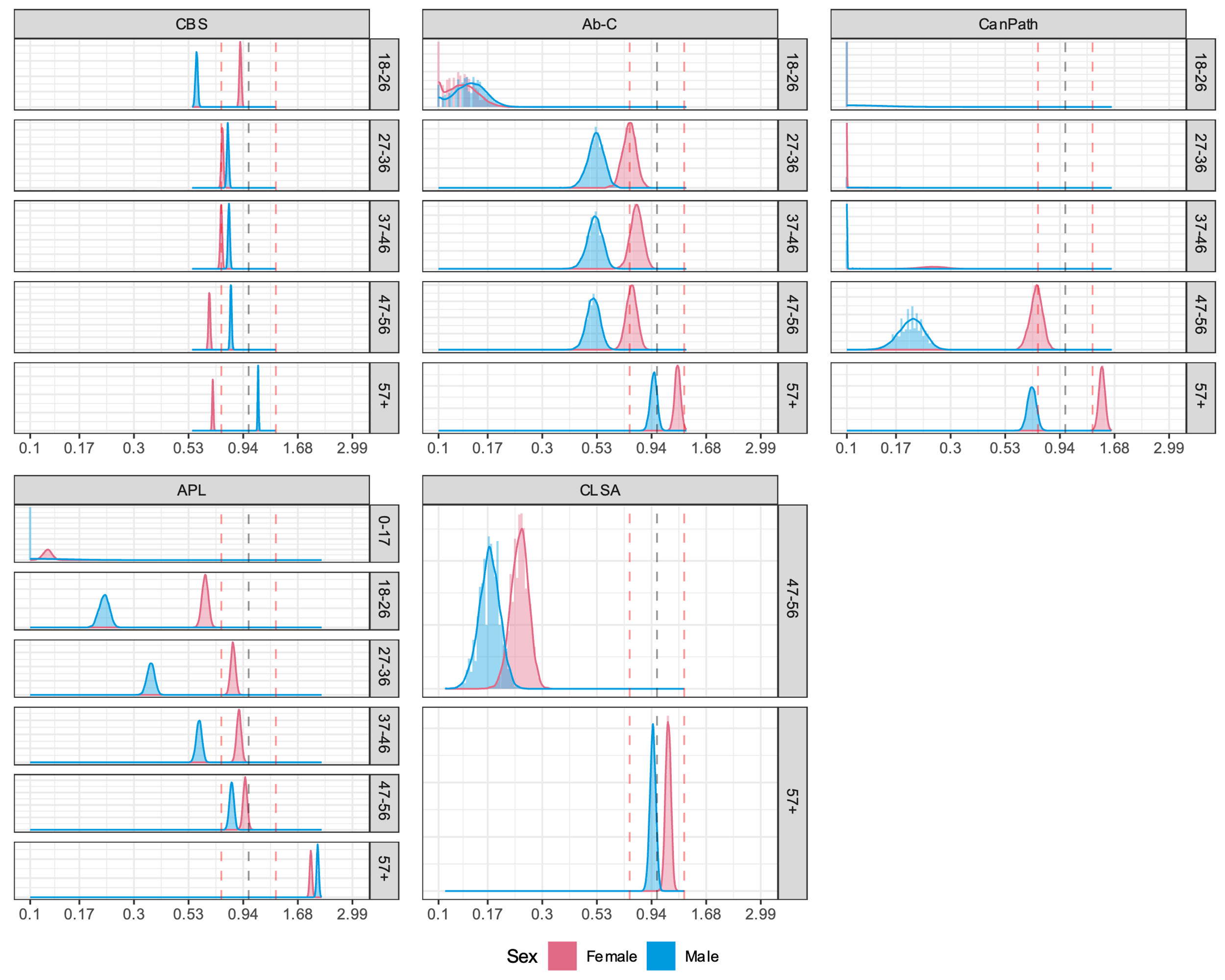

**Supplementary Figure S11: Bootstrap distribution of log10-transformed representation ratios stratified by age group, sex, and rural residence.** Distributions were generated using 5000 bootstrap resamples and plotted with a binwidth of 0.01. Representation ratios between 0.00 – 0.09 were pseudo-adjusted to a value of 0.1 prior to transformation for visualization purposes. Representativeness was calculated by dividing the proportion of study specimens collected from a subgroup by the proportion of population in the subgroup. Total population counts were estimated using the 2016 Canadian census [19]. Dashed red lines indicate representation ratio values of 0.75 and 1.33.

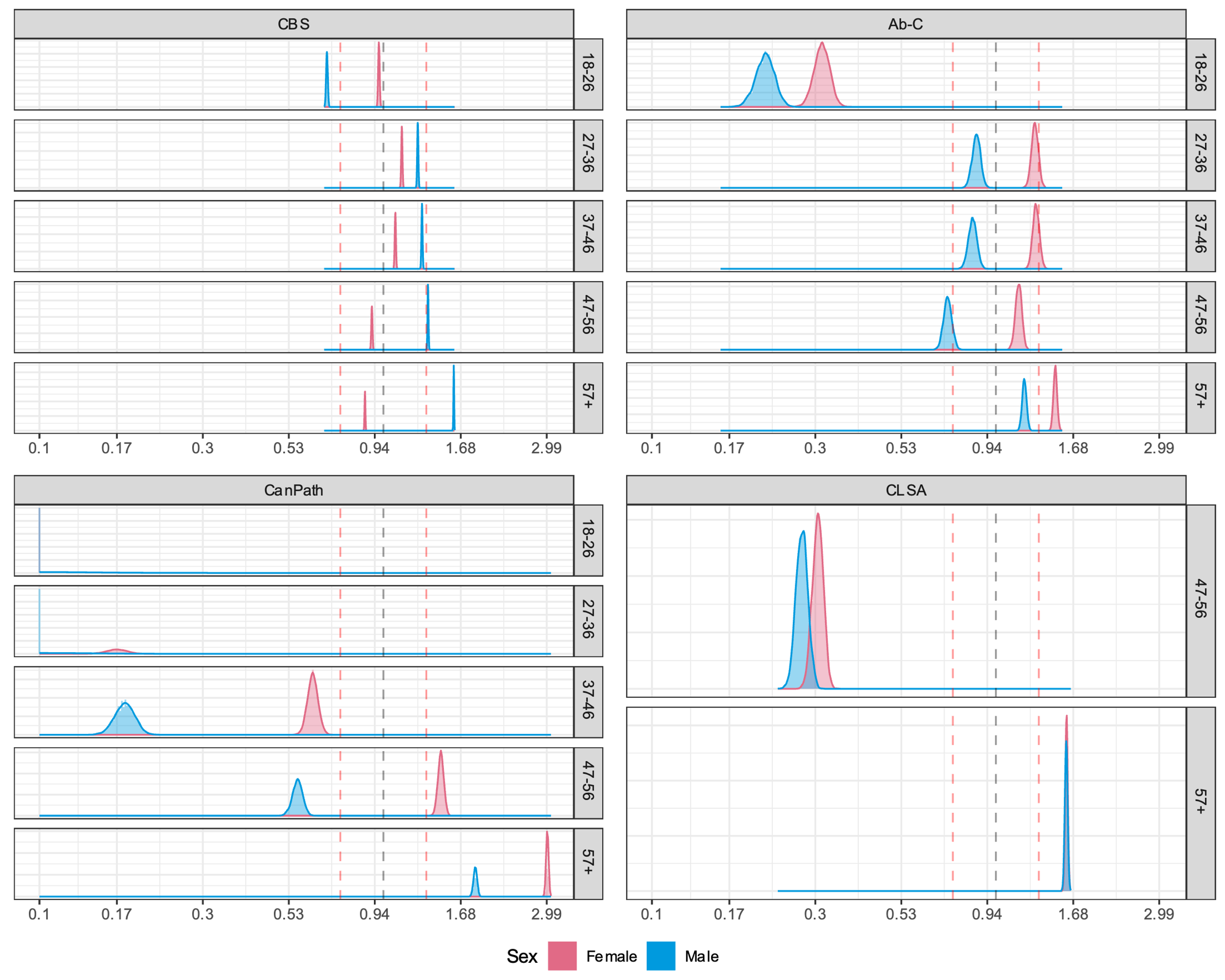

**Supplementary Figure S12: Bootstrap distribution of log10-transformed representation ratios stratified by age group, sex, and white race/ethnicity.** Distributions were generated using 5000 bootstrap resamples and plotted with a binwidth of 0.005. Representation ratios between 0.00 – 0.09 were pseudo-adjusted to a value of 0.1 prior to transformation for visualization purposes. Representativeness was calculated by dividing the proportion of study specimens collected from a subgroup by the proportion of population in the subgroup. Total population counts were estimated using the 2016 Canadian census [19]. Dashed red lines indicate representation ratio values of 0.75 and 1.33.

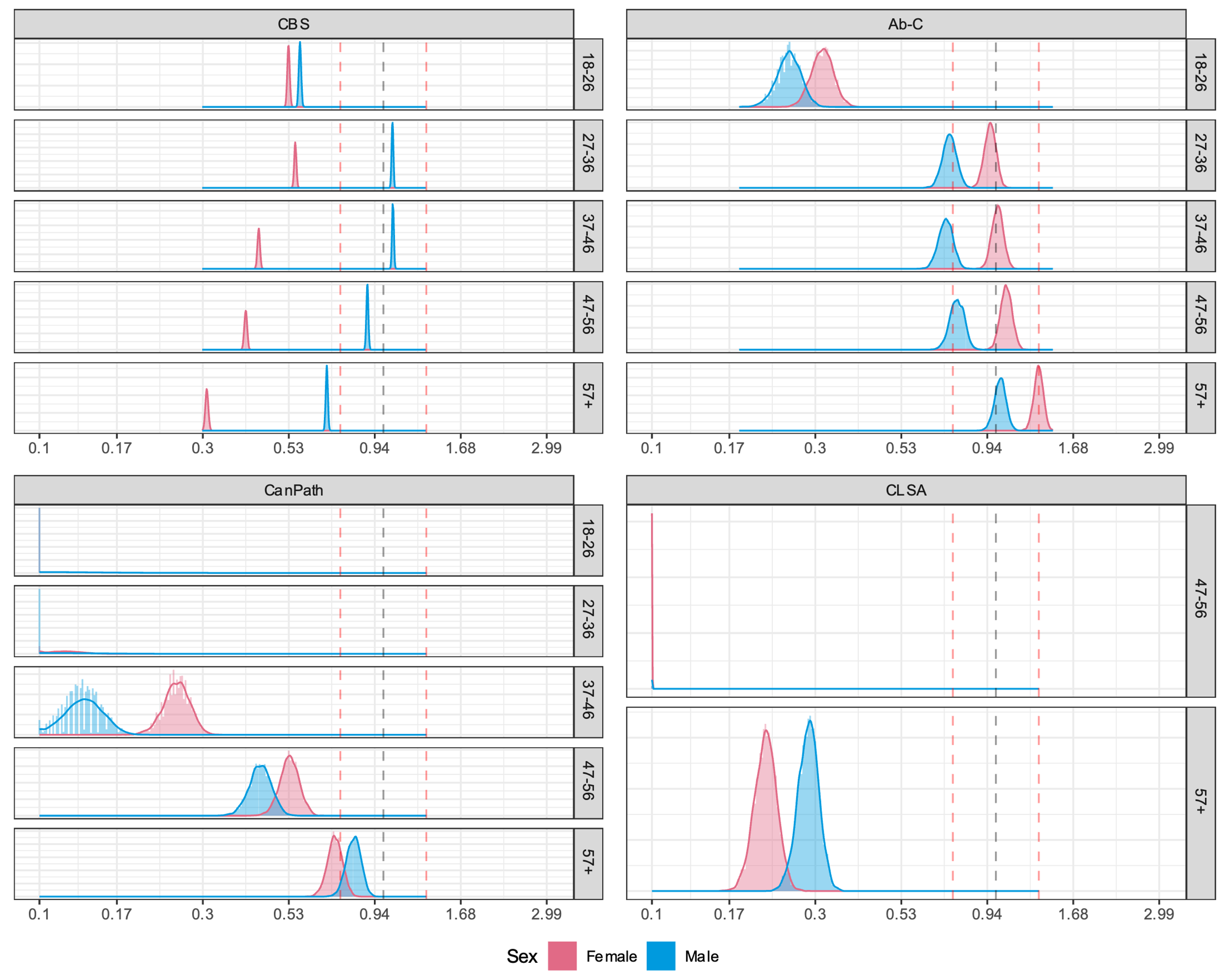

**Supplementary Figure S13: Bootstrap distribution of log10-transformed representation ratios stratified by age group, sex, and racialized minority race/ethnicity.** Distributions were generated using 5000 bootstrap resamples and plotted with a binwidth of 0.005. Representation ratios between 0.00 – 0.09 were pseudo-adjusted to a value of 0.1 prior to transformation for visualization purposes. Representativeness was calculated by dividing the proportion of study specimens collected from a subgroup by the proportion of population in the subgroup. Total population counts were estimated using the 2016 Canadian census [19]. Dashed red lines indicate representation ratio values of 0.75 and 1.33.

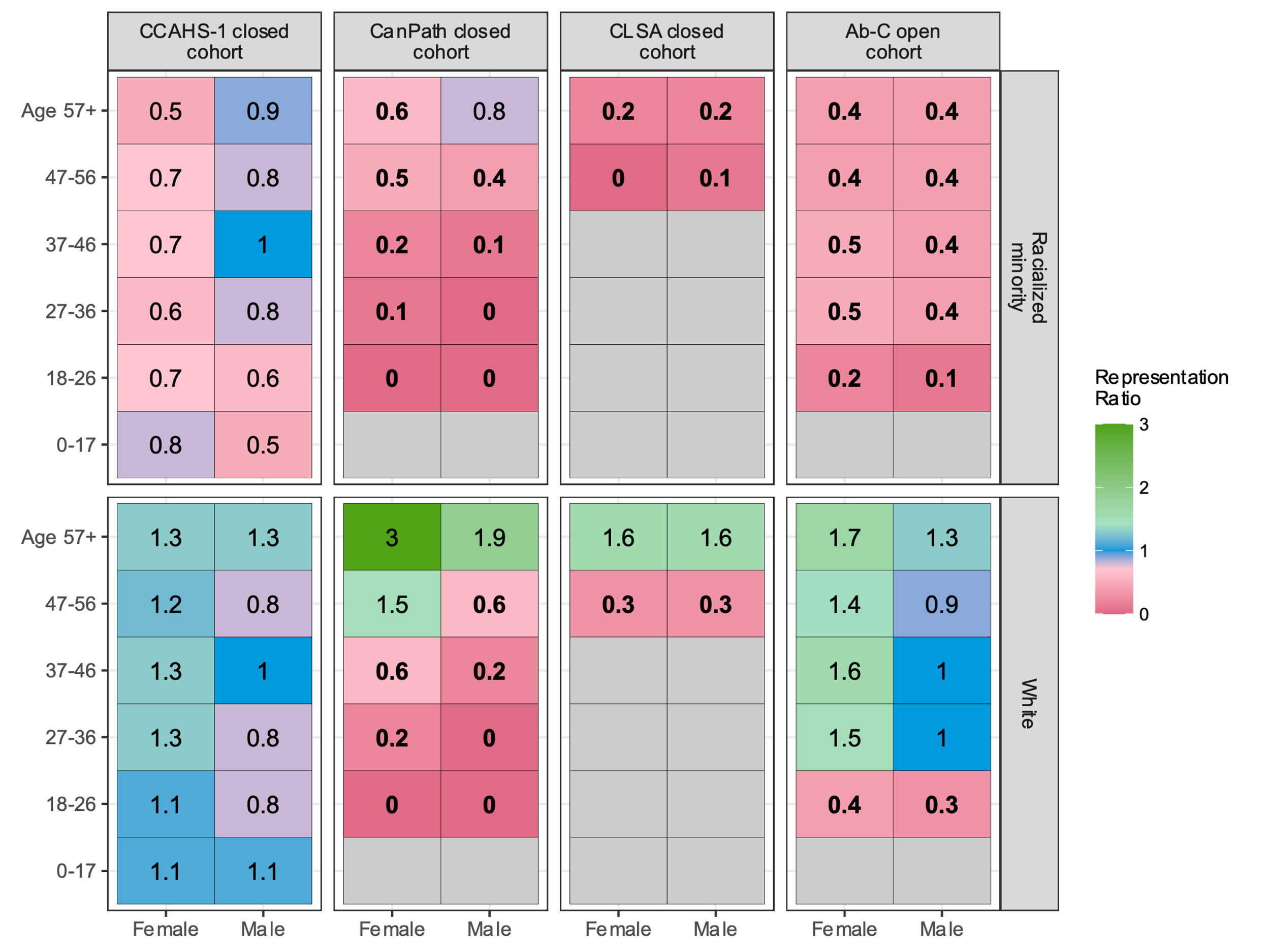

**Supplementary Figure S14: Sensitivity analysis evaluating the effect of race/ethnicity classification on SARS-CoV-2 serological study representativeness.** Participants identifying as mixed race/ethnicity were classified as white (in primary analysis they were classified as racialized minority). Representativeness was assessed by age group, sex, and racial/ethnic identity and was calculated by dividing the proportion of study specimens collected from a subgroup by the proportion of population in the subgroup. Total population counts were estimated using the 2016 Canadian census [19]. Bolded representation ratios indicate greater than 95% of subgroup bootstrap replicates produced representation ratios below 0.75. Bootstrapping was not performed for studies with weighted counts (CCAHS-1).

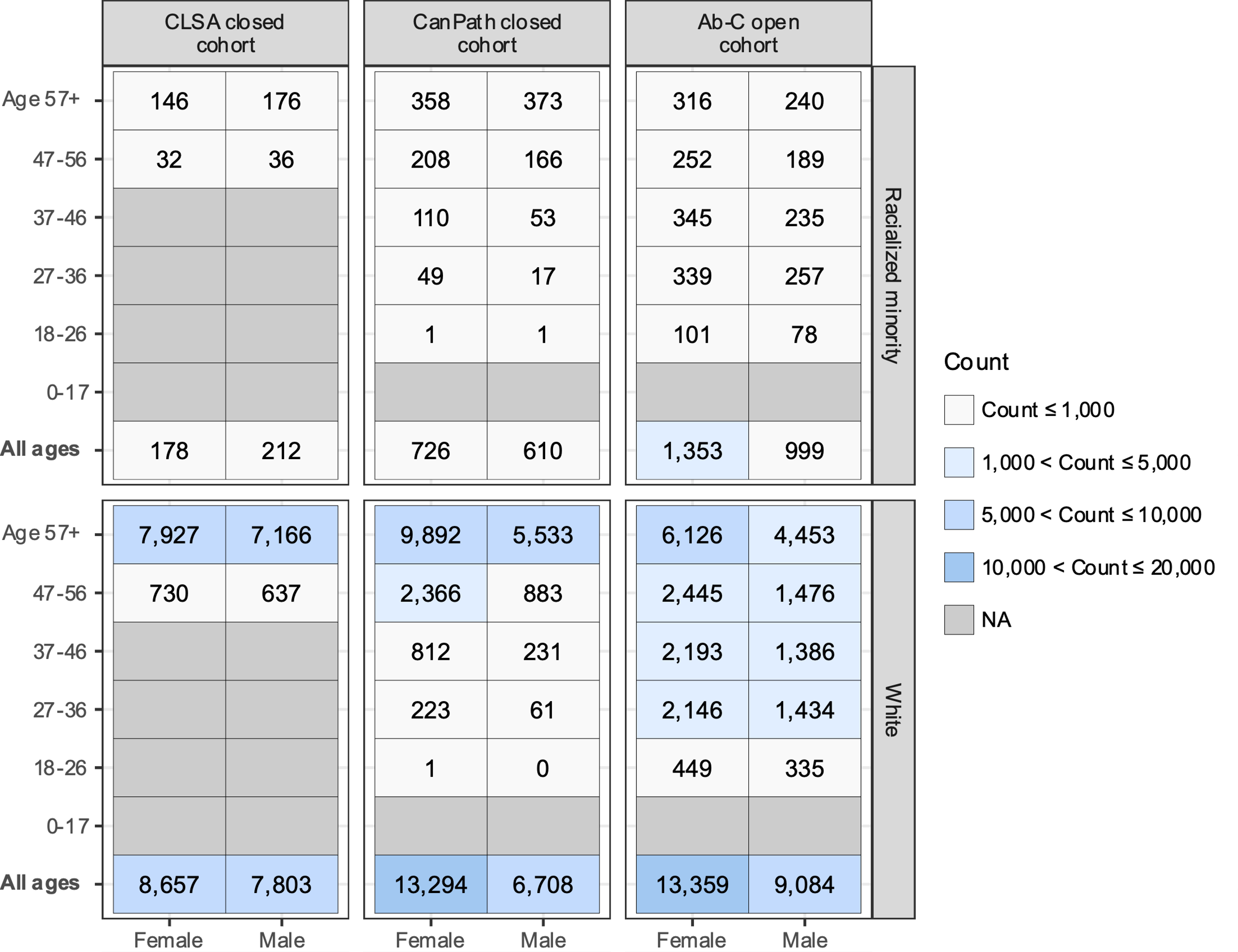

**Supplementary Figure S15: Sensitivity analysis evaluating the effect of race/ethnicity classification on SARS-CoV-2 serological study demographic composition.** Participants identifying as mixed race/ethnicity were classified as white (in primary analysis they were classified as racialized minority). Counts were calculated as the number of serological specimens contributed by each study subgroup. CCAHS-1 counts were not included due to privacy regulations.

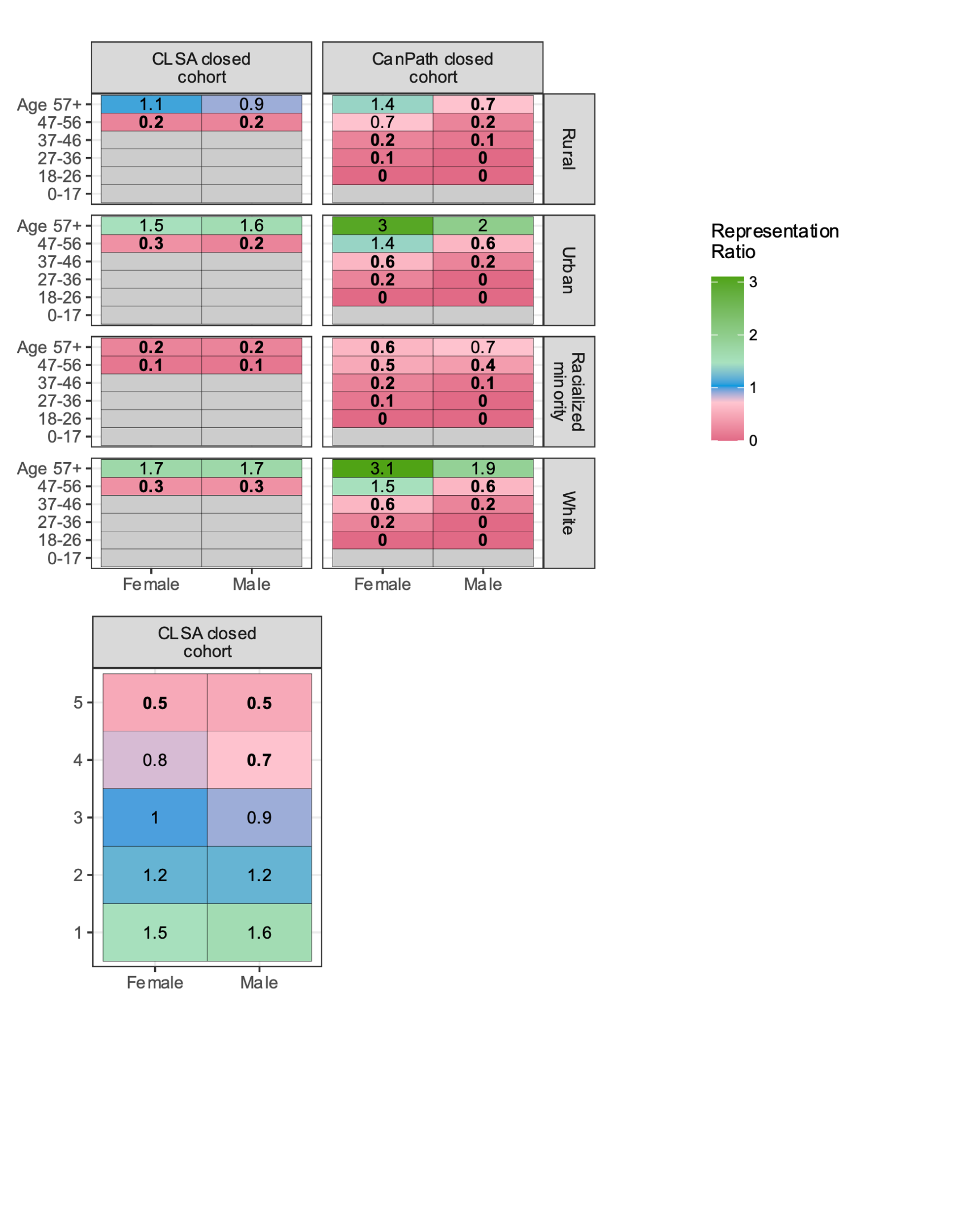

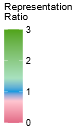

**Supplementary Figure S16: Sensitivity analysis evaluating the effect of including Indigenous-identifying census respondents on CLSA and CanPath representativeness.** Total population counts were estimated using the 2016 Canadian census and included individuals who identified as Indigenous (in primary analysis census counts excluded Indigenous-identifying individuals for CLSA and CanPath) [19]. Representativeness was assessed by age group, sex, urbanicity, racial/ethnic identity, and material deprivation quintile. Representativeness was calculated by dividing the proportion of study specimens collected from a subgroup by the proportion of population in the subgroup. Material deprivation scores were not available for the CanPath study. Bolded representation ratios indicate greater than 95% of subgroup bootstrap replicates produced representation ratios below 0.75.

**SUPPLEMENTAL TABLES**

**Supplementary table S1**: Classification table of SARS-CoV-2 serology study participant self-reported racial/ethnic identities.

|  | | **Response classification** | | |
| --- | --- | --- | --- | --- |
| **Study** | **Race/ethnicity definition** | **White** | **Racialized minority** | **Removed** |
| CBS | Ethnicity | White | Aboriginal, Asian, Other | Missing |
| CCAHS-1 | Are you X? | White | Arab, Black, Chinese, Filipino, Indigenous, Japanese, Korean, Latin American, Other, South Asian, Southeast Asian, West Asian | NA |
| Ab-C | Ethnicity | English / Irish / Scottish, French, Other European (e.g. German, Russian, Italian, Norwegian, etc.) | African (e.g. South African, Ethiopian, Nigerian, etc.), Caribbean (e.g. Jamaica, Cuba, Trinidad), Central or South American, Chinese, Filipino, Indigenous / First Nations / Inuit / Metis, Middle Eastern / Central Asian (e.g. Lebanese, Iranian, Turkish, Syrian, etc.), Oceania (e.g., Australian, New Zealander, Fijian, etc.), Other (Please specify), Other Asian (Vietnamese, Korean, Japanese, etc.), South Asian (Indian, Pakistani, Sri Lankan, etc.) | NA, Rather Not Say |
| CanPath | Race/ethnicity | White | Arab, Black, Chinese, Filipino, Japanese, Korean, Latin American / Hispanic, South Asian, Southeast Asian, West Asian, Other, Other (Specify) | Rather not say |
| CLSA | Ethnicity | White | Arab, Black, Chinese, Filipino, Japanese, Korean, Latin American, Other (Please specify), South Asian, Southeast Asian, West Asian | NA, Rather Not Say, Refused |

*Notes:* Participants who identified as non-white in race/ethnicity, or who identified as Indigenous, were classified as a racialized minority. Ambiguous or missing responses were removed for all analyses that included race/ethnicity as a stratum. NA: Not applicable.

**Supplementary table S2**: Classification table of self-reported racial/ethnic identities provided by text response in CLSA and CanPath studies.

| **Response classification** | |
| --- | --- |
| **White** | **Racialized minority** |
| Responses which included “White” or “Caucasian”. | Responses which included “Caribbean”, “Indian”,  “Indo-Caribbean”, “Asian“, “African”, “South American”, “Iraqi”, “Ugandan”, “Trinidadian”, “Chinese”, “Japanese”, “Armenian”, “Black”, “Guyanese”, “Latin-American”, “Khoisan”, “Hispanic”, “Persian”, “Assyrian”,  “Latino”, “Afro-Latino American”, “Berbere”, “Amazigh”, “Maghrebine”, “Moroccan Jewish”, “Arab”, “Afrikaans”, “Jamaican”, “Indo-European”, “Colombian”, “Middle East”, “Cherokee”, “Zimbawayan”, “Indonesian”, “Punjabi”, “Cambodian”, “Maurician”, “Taiwanese”, “Afghan”, “Tunisian”, “Mexican”, “Metis”, “Aboriginal”, or “Indigenous”. |

*Notes:* Participants who identified as non-white in race/ethnicity, or who identified as Indigenous, were classified as a racialized minority. Ambiguous or missing responses were removed for all analyses which included race/ethnicity as a stratum.

**Supplementary table S3**: Summary of demographic characteristics by SARS-CoV-2 serology study.

|  | **CBS blood donor**  **(n = 1035580 )** | | **APL outpatient laboratory**  **(n = 210905)** | | **CCAHS-1 closed cohort**  **(n = 11050)** | | **CLSA closed cohort**  **(n = 17310)** | | **CanPath closed cohort**  **(n = 21720)** | | **Ab-C open cohort**  **(n = 25110)** | |
| --- | --- | --- | --- | --- | --- | --- | --- | --- | --- | --- | --- | --- |
|  | n | % | n | % | n | % | n | % | n | % | n | % |
| **Age Group (%)** |  | | | | | | | | | | | |
| 0-17 | 0 | 0.0 | 7111 | 3.4 | 1750 | 15.8 | 0 | 0.0 | 0 | 0.0 | 0 | 0.0 |
| 18-26 | 109542 | 10.6 | 11763 | 5.6 | 1300 | 11.8 | 0 | 0.0 | 3 | 0.0 | 991 | 3.9 |
| 27-36 | 183368 | 17.7 | 26354 | 12.5 | 1200 | 10.9 | 0 | 0.0 | 355 | 1.6 | 4244 | 16.9 |
| 37-46 | 176478 | 17.0 | 27646 | 13.1 | 1450 | 13.1 | 0 | 0.0 | 1226 | 5.6 | 4219 | 16.8 |
| 47-56 | 192898 | 18.6 | 30443 | 14.4 | 1550 | 14.0 | 1485 | 8.6 | 3687 | 17.0 | 4396 | 17.5 |
| 57+ | 373294 | 36.0 | 107588 | 51.0 | 3800 | 34.4 | 15825 | 91.4 | 16449 | 75.7 | 11260 | 44.8 |
| **Material deprivation quintile (%)** |  | | | | | | | | | | | |
| 1 | 267483 | 25.8 | 54152 | 25.7 | 2550 | 23.1 | 5345 | 30.9 | 0 | 0.0 | 0 | 0.0 |
| 2 | 225349 | 21.8 | 38727 | 18.4 | 2250 | 20.4 | 3884 | 22.4 | 0 | 0.0 | 0 | 0.0 |
| 3 | 188746 | 18.2 | 32684 | 15.5 | 1950 | 17.6 | 3132 | 18.1 | 0 | 0.0 | 0 | 0.0 |
| 4 | 145317 | 14.0 | 26976 | 12.8 | 1850 | 16.7 | 2404 | 13.9 | 0 | 0.0 | 0 | 0.0 |
| 5 | 85043 | 8.2 | 17743 | 8.4 | 1450 | 13.1 | 1659 | 9.6 | 0 | 0.0 | 0 | 0.0 |
| Missing | 123642 | 11.9 | 40623 | 19.3 | 950 | 8.6 | 886 | 5.1 | 21720 | 100.0 | 25110 | 100.0 |
| **Social deprivation quintile (%)** |  | | | | | | | | | | | |
| 1 | 197381 | 19.1 | 37530 | 17.8 | 1600 | 14.5 | 2822 | 16.3 | 0 | 0.0 | 0 | 0.0 |
| 2 | 194957 | 18.8 | 28600 | 13.6 | 2100 | 19.0 | 3432 | 19.8 | 0 | 0.0 | 0 | 0.0 |
| 3 | 182655 | 17.6 | 35489 | 16.8 | 2250 | 20.4 | 3479 | 20.1 | 0 | 0.0 | 0 | 0.0 |
| 4 | 167547 | 16.2 | 33994 | 16.1 | 2200 | 19.9 | 3426 | 19.8 | 0 | 0.0 | 0 | 0.0 |
| 5 | 169398 | 16.4 | 34669 | 16.4 | 1950 | 17.6 | 3265 | 18.9 | 0 | 0.0 | 0 | 0.0 |
| Missing | 123642 | 11.9 | 40623 | 19.3 | 950 | 8.6 | 886 | 5.1 | 21720 | 100.0 | 25110 | 100.0 |
| **Race/ethnicity (%)** |  | | | | | | | | | | | |
| Racialized minority | 189304 | 18.3 | 0 | 0.0 | 1400 | 12.7 | 461 | 2.7 | 1509 | 6.9 | 5296 | 21.1 |
| White | 846276 | 81.7 | 0 | 0.0 | 9200 | 83.3 | 16389 | 94.7 | 19829 | 91.3 | 19633 | 78.2 |
| Missing | 0 | 0.0 | 210905 | 100.0 | 450 | 4.1 | 460 | 2.7 | 382 | 1.8 | 181 | 0.7 |
| **Sex (%)** |  | | | | | | | | | | | |
| Female | 433366 | 41.8 | 119362 | 56.6 | 6150 | 55.7 | 9045 | 52.3 | 14258 | 65.6 | 14815 | 59.3 |
| Male | 602214 | 58.2 | 91530 | 43.4 | 4900 | 44.3 | 8265 | 47.7 | 7462 | 34.4 | 10157 | 40.7 |
| Missing | 0 | 0.0 | 13 | (0.0 | 0 | 0.0 | 0 | 0.0 | 0 | 0.0 | 0 | 0.0 |
| **Urbanicity (%)** |  | | | | | | | | | | | |
| Rural | 133393 | 12.9 | 26045 | 12.3 | 1950 | 17.6 | 2429 | 14.0 | 1870 | 8.6 | 3156 | 12.6 |
| Urban | 902180 | 87.1 | 184859 | 87.7 | 9050 | 81.9 | 14881 | 86.0 | 19850 | 91.4 | 21923 | 87.4 |
| Missing | 7 | 0.0 | 1 | 0.0 | 0 | 0.0 | 0 | 0.0 | 0 | 0.0 | 0 | 0.0 |

*Notes:* Scores of 1 and 5 indicate the lowest and highest quantiles of deprivation, respectively. Raw CCAHS-1 counts were rounded to base 2000 according to data usage guidelines.

**Supplementary table S4**: Sensitivity analysis for percentage of SARS-CoV-2 study demographic subgroups with greater than 25 collected specimens.

|  | | Demographic subgroups | | | | |
| --- | --- | --- | --- | --- | --- | --- |
| Study  (specimen count) | Months sampled | | Age, Sex, Province, Month | Age, Sex, Province, Urban, Month | Age, Sex, Province, Race/Ethnicity, Month | Age, Sex, Province, Race/Ethnicity, Urban, Month |
| CBS blood donor (1,035,580) | 41 | | 92% | 74% | 70% | 52% |
| APL outpatient laboratory (210,905) | 27 | | 94% | 84% | NA | NA |
| Ab-C open cohort (25,110) | 18 | | 31% | 20% | 21% | 14% |
| CanPath closed cohort (21,720) | 10 | | 40% | 31% | 33% | 27% |
| CLSA closed cohort (17,310) | 11 | | 50% | 36% | 34% | 27% |
| CCAHS-1 closed cohort (11,050) | 6 | | 33% | 20% | 21% | 13% |

*Notes:* Study participants identifying as mixed race/ethnicity (white and racialized minority) were classified as white. Date of sample collection was binned into 2-month intervals. All specimen counts were unweighted.
